# Beyond Binary Diagnosis: A Genetic Algorithm Optimized Hybrid Machine Learning Model for PCOS Phenotype Prediction

**DOI:** 10.64898/2026.10.01.26364554

**Authors:** Priyanka Roy, Snehal Dnyane, Komal Sakhale, Saurav Mallik

## Abstract

In this work, we investigate the homogeneity of performance in binary classification between the Rotterdam phenotypes and phenotype prediction. For this purpose, a multi-task neural network architecture is designed to improve the prediction accuracy of phenotypes while maintaining the accuracy of binary classification. Using a clinical PCOS dataset which contains unlabeled samples of patients that cannot be attributed to any of the known phenotypes, three models are evaluated: optimized binary Random Forest (RF), six-class weighted class RF, and multi-task neural network (with a common hidden layer and two heads for binary and six-class classifications) optimized by a genetic algorithm. Techniques used for evaluation include ten-fold out-of-fold predictions, paired McNemar tests, bootstrap tests, ablation study, and clustering (Adjusted Rand Index, ARI). Binary RF model reached 90.2% accuracy; however, recall of different phenotypes was significantly unbalanced – from 75.0% of phenotype B to 97.8% of phenotype A. This difference of 22.8 % points was observed (23.3 points) even after excluding phenotype-specific features. While direct phenotype prediction improved recall for common phenotypes, it decreased recall for phenotype B to 43.8% and unclassified cases to 31.5%. Our multi-task model preserved 90.2% binary accuracy and significantly recovered phenotype B recall to 87.5% and unclassified cases to 79.6% (p<0.05). The cluster analysis confirmed the weak correlation between the results and labels (ARI=0.045). The ablation analysis demonstrates that feature selection and class weighting drive this improvement, isolating the specific components responsible for performance gains. We establish that aggregate binary accuracy does not reflect the heterogeneous detection of PCOS phenotypes. Direct phenotype prediction alone redistributes the classification errors, but the multi-task architecture successfully improves the detection of rare phenotypes without compromising binary accuracy. Phenotype-disaggregated evaluations, supported by significance testing and ablation, are essential for clinical classifier research.

## 1. Introduction

Polycystic Ovary Syndrome (PCOS) is one of the most frequent hormonal disorders among reproductive-age women. Women suffering from PCOS may have problems with ovulation, including irregular and lack of ovulation cycles. Other PCOS symptoms include increased androgen production, which can manifest itself as hirsutism, skin acne, and alopecia. At the same time, some women may have polycystic ovaries on ultrasound imaging but do not have hormonal symptoms. Taking this diversity into account, the Rotterdam ESHRE/ASRM Consensus Group created a diagnostic criterion for PCOS in 2004 – a woman has PCOS if she has two out of three possible symptoms: irregular ovulation, hyperandrogenism, and polycystic ovarian morphology. Having three possible symptoms and needing only two to meet the criteria means that there are four possible phenotypes of PCOS A through D. Clinicians consider these phenotypes quite different in terms of presentation and long-term health consequences.

Machine learning research on PCOS frequently overlooks these clinical distinctions. There is one commonly used dataset for this type of research, consisting of 541 PCOS patients gathered in ten hospitals in Kerala, India and uploaded to Kaggle. Most studies on PCOS classification use this dataset to train a machine learning model that answers a simple binary question: does this patient have PCOS or not? Reported accuracy for such models varies between 80% and nearly 99%, with the use of methods ranging from standard logistic regression to complex ensembles of tree-based models with feature selection and optimization. Some studies employ hybrid models combining neural network-based and tree-based classification and optimization techniques like genetic algorithms. Crucially, almost none of these studies examine whether the reported accuracy is consistent across the four distinct phenotypes of PCOS.

Predictive performance consistency is directly analyzed in this study. Well-calibrated Random Forest classifier achieves 90.2% accuracy on this dataset, consistent with earlier results. Phenotype-aggregated results show that the same classifier reaches near-perfect accuracy when predicting one of the phenotypes (97.8%) but performs much worse when predicting the rarest one (75.0%). This significant difference is hidden behind the overall accuracy measure. The difference is preserved when all the variables used for defining phenotypes are discarded, which means that it could not be caused by the fact that one of the groups was built to be easier to detect than another. Predicting the phenotype directly, instead of predicting its binary label, does not solve the problem either. On the contrary, it moves the difference in a different direction: detection of the common phenotype becomes 100% accurate, while detection of the rare phenotype drops down to 43.8%.

The main technique employed in this paper deals with this issue. Specifically, we introduce a tree-based feature ranking stage that feeds inputs into a multi-task neural network optimized using our customized genetic algorithm. The network simultaneously predicts binary labels and phenotypes from a common representation. For this particular dataset, this technique makes the recall of the rare phenotype equal to 87.5%, and makes detection of the most difficult, most ambiguous PCOS subtype increase from 31.5% to 79.6%. At the same time, the overall accuracy remains at 90.2%. Both improvements are confirmed using a significance test (Section 3.8). Next, an ablation study is conducted to find out what contributes to the improvements (Section 3.8). This paper advocates for phenotype-disaggregated evaluation based on significance tests and ablation studies instead of aggregated accuracy measures and inference.

### 1.1 Literature Survey

The first cluster of studies addresses prediction of PCOS at the population level, using broad sources of data rather than clinical datasets, on which most classifiers operate. Joo et al. (2020) [1] combined polygenic risk scores with a phenome-wide association study on more than one hundred thousand electronic health records. They discovered biological connections between PCOS and other diseases, such as obesity and type 2 diabetes, instead of creating a classic diagnostic classifier. Zad et al. (2024) [5] followed a similar approach. They trained logistic regression, SVM, gradient-boosted trees, and random forests on more than thirty thousand outpatient records. Importantly, they developed their labels directly from PCOS components derived from the Rotterdam criteria, extracted from the EHR. This makes this paper one of the few in this field that touches the diagnostic framework, even though the models themselves produce one score per patient, rather than a PCOS phenotype. A number of review papers (Memon et al., 2025 [2]; Ghaderzadeh et al., 2025 [4]; Govindharajan et al., 2025 [7]; Joyti et al., 2026 [10]) examine this field and reach consistent conclusions – namely, that classical and ensemble methods dominate the field of structured clinical data analysis, while deep learning performs better in imaging. Both fields face persistent issues, such as small sample size, limited external validation, and near-complete absence of subgroup-level reports.

The second cluster is closer to the structured clinical dataset used in this paper and is centered around binary PCOS detection. Nasim et al. (2022) [6] proposed a feature selection method using chi-squared criterion along with ten tuned classifiers and reported a Gaussian Naive Bayes model achieving a perfect accuracy on their clinical dataset. Rahman et al. (2024) [9] used Mutual Information for feature selection on the same 541-patient Kaggle dataset that serves as a basis for this paper. They discovered that both AdaBoost and Random Forest reach an accuracy of around 94% and even created a deployable web tool for their model. Agirsoy and Oehlschlaeger (2025) [8] take a diagnostics-oriented path by dividing their features into three categories according to the Rotterdam criteria – clinical, biochemical, and ultrasound – and testing which combinations perform best with XGBoost, achieving almost a perfect result with external validation. Silva et al. (2022) [3] comes closest to the topic of this paper – they select features using BorutaShap and Random Forest and then use those to divide their PCOS-positive patients into two phenotypic clusters based on metabolic markers, explicitly stating that PCOS patients are not homogeneous. Yet the authors did not test the performance of their diagnostic classifier on each cluster, as it is done in this paper.

The third cluster of papers is methodologically closest to the model used in this paper. These studies implement hybrid architectures with optimization, aiming for high accuracy on a single binary or image-level classification task. Ghosh and Srinivasan (2025) [12] optimized an ensemble of EfficientNetB7 and DenseNet201 using a genetic algorithm, reaching over 99% accuracy on ultrasound images, and are among the few PCOS studies to apply a genetic algorithm to hyperparameter tuning rather than grid or random search. Faris et al. (2025) [13] take a similar approach on structured data, combining k-means clustering with genetic feature selection to build a bagging ensemble on the same 541-patient dataset used throughout this paper. Pallavi and Soumya (2026) [11] extend the hybridization idea further still, combining a Bi-GAN with a physics-informed neural network across ultrasound, hormonal, and metabolic inputs for cloud-edge deployment. What unites all three is that, despite the architectural complexity, the goal remains near-perfect classification into PCOS-positive and PCOS-negative groups; none checks whether that improvement holds evenly across the phenotypic subgroups Silva et al. (2022) [3] identified. This is precisely the gap this paper addresses.

The reviewed studies provide examples of three independent approaches to predicting PCOS, given in **Table 1**: population-level studies of genetic and EHR data to stratify risk, but not diagnose patients [1], [5], classifier research on structured clinical datasets treating PCOS as a binary outcome [3], [6], [8], [9], and hybrid optimization-tuned models, pushing accuracy as high as possible, but still predicting only a single binary label [11], [12], [13]. The recent review papers in this field [2], [4], [7], [10] agree on the same limitations of small samples, weak external validation, and no subgroup-level reporting. Only one paper [3] goes as far as to cluster the PCOS-positive patients into phenotypes, yet does not test whether performance of the classifier holds in each cluster. None of the reviewed studies combine an optimization-tuned hybrid model with phenotype-based evaluation. This is the gap addressed in this paper.

**Table 1:** Summarized Literature Survey.

| Year | Study | Binary PCOS Classification | Hybrid / GA-Optimized | Phenotype-Level Evaluation | External / Real-World Clinical Data |
| --- | --- | --- | --- | --- | --- |
| 2020 | Joo et al. [1] | No | No | No | Yes |
| 2022 | Silva et al. [3] | No | No | No | No |
| 2022 | Nasim et al. [6] | Yes | No | No | No |
| 2024 | Zad et al. [5] | Yes | No | No | Yes |
| 2024 | Rahman et al. [9] | Yes | No | No | No |
| 2024 | Kermanshahchi et al. [17] | Yes | No | No | No |
| 2025 | Agirsoy & Oehlschlaeger [8] | Yes | No | No | Yes |
| 2025 | Ghosh & Srinivasan [12] | Yes | Yes | No | No |
| 2025 | Faris et al. [13] | Yes | Yes | No | No |
| 2026 | Pallavi & Soumya [11] | Yes | Yes | No | No |
| 2026 | Tasnim et al. [15] | Yes | Yes | No | No |
| 2026 | Smithakranthi & Haritha [16] | Yes | Yes | No | No |
| **2026** | **This Work (Ours)** | **Yes** | **Yes** | **Yes** | **Partial\*** |

The sixteen reviewed studies exemplify three distinct approaches to PCOS prediction: population-level genetic and electronic health record analyses that stratify risk without diagnosing individual patients [1], [5]; binary classification on structured clinical datasets [3], [6], [8], [9]; and hybrid optimization-tuned models that pursue maximal binary accuracy [11], [12], [13]. Recent reviews [2], [4], [7], [10] converge on the same open challenges: small samples, limited external validation, and no subgroup-level reporting. Only one study [3] clusters PCOS-positive patients into phenotypes, yet does not test whether classifier performance is consistent across those clusters. No prior work combines a genetically optimized hybrid model with phenotype-stratified evaluation—the gap this paper addresses directly.

A terminological note and a fast-moving recent literature both bear directly on how this gap should be read today. In May 2026, a fifty-six-organization global consensus process led by Teede et al. (2026) [18] formally renamed PCOS to Polyendocrine Metabolic Ovarian Syndrome (PMOS), arguing that “polycystic ovary syndrome” misleadingly centers the ovary and obscures the condition’s endocrine and metabolic breadth. This paper retains “PCOS” and the Rotterdam phenotype labels throughout, for continuity with the dataset, the cited literature, and the diagnostic criteria actually used to construct the phenotype labels analyzed here, but every finding below should be read as applying to PMOS under the new nomenclature, and future extensions of this work should adopt the updated terminology directly.

The pace of publication on this specific problem has also not slowed. Since the literature reviewed above, at least three further hybrid or feature-fusion PCOS classifiers have appeared in 2026, none of which evaluate phenotype-level performance. Tasnim et al. (2026) [15], publishing in Health Information Science and Systems, fused ResNet-18 and GoogLeNet deep features for ovarian ultrasound classification, reaching 99.19% accuracy with a lightweight classification head; Smithakranthi and Haritha (2026) [16] combined a feature-sequence convolutional network with a gated feature-learning network on tabular clinical variables, reporting up to 97% accuracy; and Kermanshahchi et al. (2024) [17] built a machine-learning model for PCOS detection and classification directly from pelvic ultrasound. All three continue the pattern identified above: architectural sophistication - feature fusion, gated networks, ensemble deep learning - is aimed exclusively at a single binary or image-level target, with no subgroup or phenotype-level accuracy reported. If anything, the continued absence of phenotype-stratified evaluation in venues publishing throughout 2026 strengthens rather than weakens the case that this gap is a structural blind spot in the field, not a temporary lag that the most recent literature has already closed.

### 1.2 Research Gap

In reading through the three categories of previous work here, the same picture emerges regardless of the technical sophistication of the model itself. Work which operates at the population level, like that of Joo et al. (2020) [1] and Zad et al. (2024) [5], takes seriously the issue of diverse and large-scale data sets but still reduces the output to a risk score, even in cases where, as in the latter work, the initial labels themselves had been generated based on elements of the Rotterdam criteria and might very well have served as the basis for a phenotype-focused target. Work which operates directly on the clinical benchmark that forms the focus of this paper, including Nasim et al. (2022) [6], Rahman et al. (2024) [9], and Agirsoy and Oehlschlaeger (2025) [8], all of them achieve outstanding levels of accuracy in their results but without a single exception take the detection of PCOS to be a binary issue. Even Silva et al. (2022) [3], the study which best mirrors the current paper in its grouping of PCOS-positive subjects into phenotypic clusters, did nothing more than generate the clusters and not even check whether there was any significant difference in classifier accuracy between them.

The third cluster addresses only one part of the problem while leaving another largely untouched. Ghosh and Srinivasan (2025) [12], Faris et al. (2025) [13], and Pallavi and Soumya (2026) [11] bring real architectural and optimization sophistication to PCOS prediction - genetic algorithms, ensembles, transfer learning, hybrid generative and physics-informed networks - demonstrating that this field is not short on modeling ambition. That sophistication, however, has been directed entirely at improving a single binary or image-level output, with no attention to whether the improvement is shared evenly across the phenotypic subgroups the diagnosis itself comprises. Recent systematic reviews (Ghaderzadeh et al., 2025 [4]; Govindharajan et al., 2025 [7]; Joyti et al., 2026 [10]) reach a similar conclusion from a different angle, repeatedly flagging small sample sizes and limited external validation as open problems in this domain.

Collectively, this gap does not arise because there is a lack of methodology. The point is that no paper discussed in this section has taken the step to verify whether the performance demonstrated in each one still holds when the patients are categorized according to their Rotterdam phenotype, and that no model has been constructed with the intention of ensuring that the rarest phenotypic group is not used as collateral damage for improving overall performance. This is precisely the gap this paper addresses directly.

### 1.3 Contribution

The goal of this research is two-fold. The first is to design, develop, and optimize a hybrid machine learning architecture for PCOS diagnosis: a Random-Forest-guided feature ranking step feeding a custom multi-task neural network, with every network parameter - hidden layer width, learning rate, the balance between binary and phenotype loss terms, L2 regularization strength, the number of Random-Forest-ranked features retained, and training length - tuned by a genetic algorithm built specifically for this problem rather than grid search, random search, or manual tuning. The second goal is diagnostic: using this optimized model to test whether a classifier can identify a patient’s Rotterdam phenotype, not just whether she has PCOS, without sacrificing performance on the rarest phenotype. These goals are interdependent: as shown in Section 3, a plain multi-class classifier trained on the same features without genetic-algorithm optimization actively degrades rare-phenotype performance.

Four contributions follow directly from the gaps identified above. The first is the construction of proxy Rotterdam phenotype labels for the PCOS benchmark dataset, explicit about where the available features fall short of the clinical gold standard, with patients who do not clearly satisfy the diagnostic rule reported as a distinct category rather than forced into one of four phenotypes for convenience. The second is the finding that a properly tuned binary classifier’s accuracy is not uniform across Rotterdam phenotypes, confirmed through a direct circularity check ruling out the possibility that this is simply an artifact of how the phenotype labels were constructed. The third is the demonstration that predicting phenotype directly, rather than checking it after binary training, is not a straightforward improvement but a trade-off that preserves accuracy on common phenotypes at the cost of rare ones - a direct comparison missing in prior hybrid or optimization-driven PCOS studies. The fourth is a hybrid multi-task model, implemented independently rather than assembled from existing components, that recovers rare-phenotype performance with statistically confirmed improvement over the multi-class model while preserving accuracy on the standard diagnostic task. The follow-up ablation (Section 3.8) further isolates the specific components driving this recovery, providing clear insights into the model’s behavior and establishing that representation sharing between the two tasks is not an absolute requirement for performance restoration.

## 2. Methodology

### 2.1 Data Preparation

The analysis employs the freely available Kerala PCOS dataset (Kottarathil, 2020, [14]) which records 44 clinical, hormonal, anthropometric, and ultrasound measurements for 541 women, 177 (32.7%) of whom have a positive PCOS diagnosis. There were two entries where a letter substituted for a number in recording a hormone measurement; both cases were changed to missing and imputed by column median. Biochemical measures of hyperandrogenism (total testosterone, free testosterone, DHEAS) and free androgen index are absent from the dataset. Consequently, all phenotypes identified in the downstream processing are based on indirect clinical measurements.

### 2.2 Rotterdam Phenotype Derivation

According to the Rotterdam ESHRE/ASRM-Sponsored PCOS Consensus Workshop Group (2004), a patient is diagnosed with PCOS if she satisfies at least two of the following three criteria: ovulatory dysfunction, hyperandrogenism, and polycystic ovarian morphology. Three binary flags were generated for each patient to identify these phenotypes. Ovulatory dysfunction was derived from menstrual cycle regularity measurements. Hyperandrogenism was approximated through a four-signal majority vote based on the presence of hirsutism and/or acne, and LH and AMH measurements above the 85th percentile. This percentile threshold was determined exclusively from non-PCOS patients to avoid self-referential definitions. Polycystic ovarian morphology was defined as the presence of 10 or more follicles in either ovary.

The phenotype classification was applied to the 177 PCOS-positive patients. Patients satisfying all three binary flags were assigned phenotype A. Ovulatory dysfunction plus hyperandrogenism corresponded to phenotype B. Hyperandrogenism and polycystic morphology indicated phenotype C. Finally, ovulatory dysfunction and polycystic morphology corresponded to phenotype D. Patients who were not satisfied with two or more of these binary flags were assigned to the Unclassified category to maintain classification integrity.

### 2.3 Circularity Control

Seven of the variables used to define the phenotypes—menstrual cycle regularity, hirsutism, acne, LH, AMH, and two follicle count variables—are available as standard features in the dataset. To prevent confounding, a second, reduced feature set was generated by removing these seven phenotype-defining variables, resulting in 34 features instead of 41. The evaluations were performed on both datasets, using each feature set independently.

### 2.4 Binary Classifier Benchmarking and Tuning

Five commonly used PCOS classifiers—logistic regression, random forest, gradient boosting, an RBF-kernel support vector machine, and XGBoost—were evaluated on the full feature set. Accuracy, ROC-AUC, F1, precision, and recall were computed for each via ten-fold cross-validation. Each classifier was tuned using RandomizedSearchCV over twenty iterations optimizing ROC-AUC, and the tuned cross-validated AUC was compared against the untuned baseline.

### 2.5 Per-Phenotype Evaluation Protocol

To obtain consistent evaluations of classifier performance, particularly given that subgroup B contains only sixteen patients, a pool of out-of-fold predictions was collected throughout the ten-fold cross-validation. This ensures each patient was predicted once by a classifier trained without their data. Accuracy and recall per phenotypic group were computed from this prediction pool. 95% confidence intervals were estimated through a 2,000-resample bootstrap to quantify true variability.

### 2.6 Multi-Class Phenotype Model

To determine if a multi-class approach addresses accuracy disparities, a class-weighted random forest classifier was trained on six classes: Control, A, B, C, D, and Unclassified. Class weights were assigned inversely proportional to class size to prevent bias towards majority classes. Performance was assessed using cross-validated macro-F1 and overall accuracy from out-of-fold predictions, with detection recall for every class compared to the original binary classifier.

### 2.7 Hybrid Multi-Task Architecture

The multi-class approach addresses the data constraints inherent in classifying six groups, where phenotype B offers only sixteen examples. The hybrid model draws on the much larger binary-task sample by allowing the two tasks to share a hidden representation.

**Figure 1** illustrates the architecture: a feature-selection module followed by a custom neural network. The first module uses a Random Forest to rank clinical features by importance, with a genetic algorithm selecting the optimal number of top-ranked features to retain. The second module is a feedforward neural network implemented in NumPy. It features a single hidden layer branching into two heads: a sigmoid head predicting the binary PCOS label, and a softmax head predicting one of six phenotype classes. Both heads are trained jointly via backpropagation with the Adam optimizer. The binary head employs binary cross-entropy loss, and the phenotype head uses a class-weighted cross-entropy loss to address phenotype imbalance. The shared hidden layer allows information from the binary task, trained on all 541 patients, to support the phenotype task.

**Figure 1:**
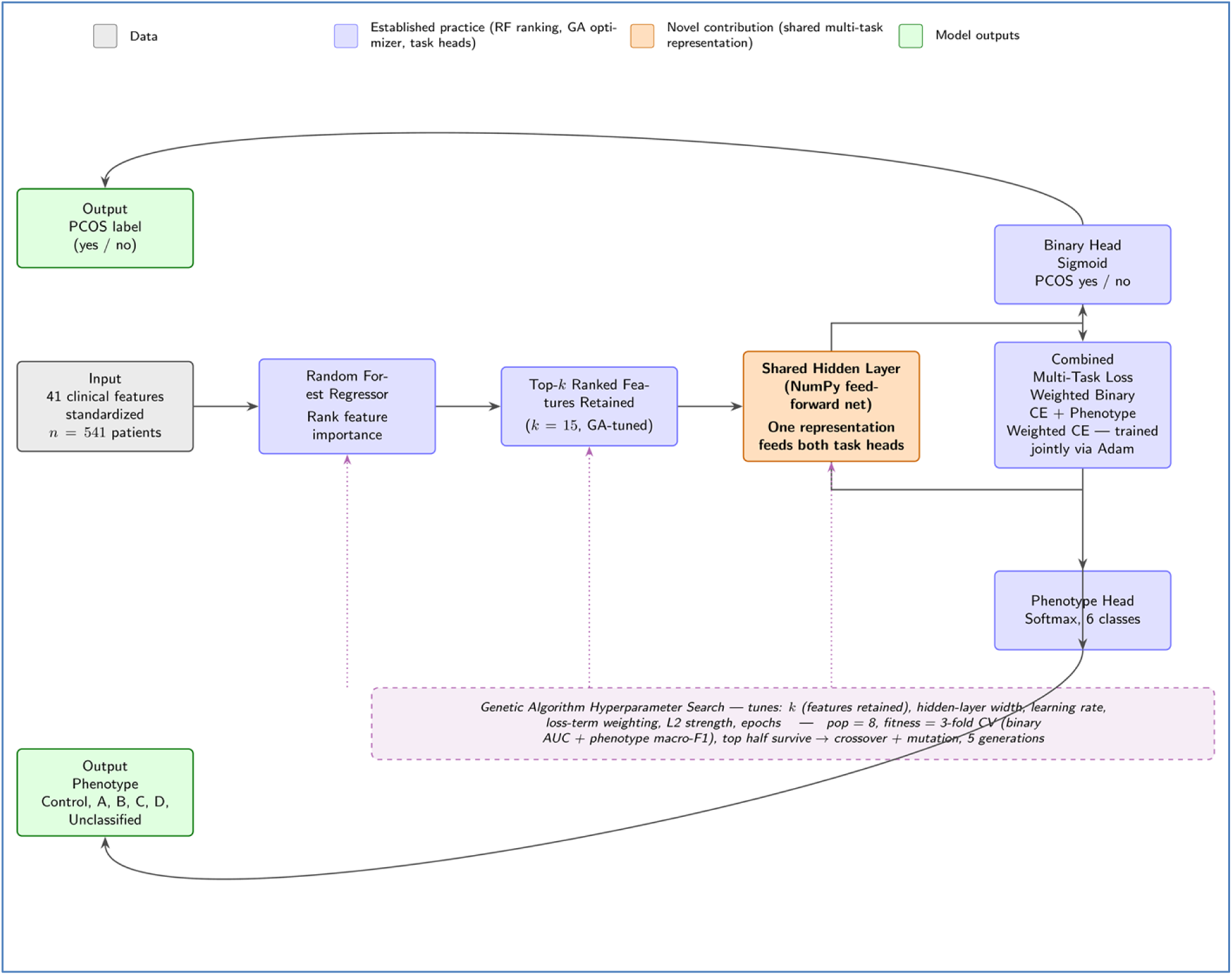
Full architecture of the proposed hybrid model. The Random Forest ranking step and genetic-algorithm-based optimization (drawn from prior practice) feed into a custom neural network where a shared hidden representation serves both the binary and phenotype classification heads through a unified training objective.

### 2.8 Genetic Algorithm Hyperparameter Optimization

The hyperparameters of the hybrid model—hidden layer size, learning rate, weight of the loss function terms relative to each other, L2 regularization strength, number of Random Forest-ranked features used, and number of training epochs—were optimized through a custom genetic algorithm. Eight initial candidates were ranked using threefold cross-validation based on a combined metric of binary AUC and phenotype macro-F1 scores. The top half of candidates survived into the next generation unaltered. Open slots in subsequent generations were filled through crossover between two survivors followed by random mutations, iterating over five generations. The single best configuration was then evaluated using the ten-fold out-of-fold protocol applied to the other models.

#### Representation

The solution encoding consists of a vector of six genes, each holding a hyperparameter value. Each gene takes a value from a predefined list (**Table 2**): hidden layer size in {8, 16, 32, 64}; learning rate in {0.01, 0.03, 0.05, 0.08}; binary/phenotype loss function weighting coefficient alpha in {0.20, 0.35, 0.50, 0.65, 0.80}; L2 regularization strength in {0, 0.001, 0.01}; number of Random Forest-ranked features kept k in {15, 25, 34, 41}; number of training epochs in {150, 250, 350}.

**Table 2:** Search space for genetic algorithm and corresponding solution found (population = 8, generations = 5, fitness = 0.5×AUC + 0.5×macro-F1, seed = 42; highest fitness achieved = 0.812)

| Hyperparameter | Candidate values searched | GA-selected value |
| --- | --- | --- |
| Hidden-layer width | 8, 16, 32, 64 | 64 |
| Learning rate | 0.01, 0.03, 0.05, 0.08 | 0.03 |
| Loss weighting ($\alpha$) | 0.20, 0.35, 0.50, 0.65, 0.80 | 0.50 |
| L2 regularization | 0, 0.001, 0.01 | 0.01 |
| Number of RF-ranked features (k) | 15, 25, 34, 41 | 15 |
| Training epochs | 150, 250, 350 | 350 |

#### Initialization

Chromosomes for the first-generation population of eight solutions are sampled independently and uniformly from their candidate sets. A fixed random seed (42) ensures full reproducibility.

#### Fitness evaluation

Each chromosome is decoded into a hybrid model configuration and evaluated using threefold cross-validation. AUC and macro-F1 scores are averaged across folds and combined into a single fitness score with equal weighting: fitness = 0.5 × mean(AUC) + 0.5 × mean(macro-F1).

#### Selection

Chromosomes are sorted in decreasing order of fitness within a generation. The fitter half survives into the next generation unchanged.

#### Crossover

The four vacated slots are filled by sampling two chromosomes randomly from the fitter half and performing uniform crossover to produce four offspring.

#### Mutation

Each gene of each offspring is independently re-sampled from its candidate values with a probability of 0.25.

#### Termination and final selection

This cycle runs for five generations, with the best chromosome across the run retained. The search reached its best configuration (fitness = 0.812) by generation 2 and held it through generation 5: a 64-unit hidden layer, learning rate 0.03, equal loss weighting (alpha = 0.5), L2 regularization of 0.01, the top 15 Random-Forest-ranked features, and 350 training epochs (**Table 2**). This configuration was then evaluated using the ten-fold cross-validation procedure.

### 2.9 Unsupervised Validation

To verify whether the phenotypes reflect an inherent structure in the feature space without relying on supervised models, K-means clustering was applied to the PCOS-positive patients across several cluster numbers. Comparisons between clusters and phenotypes used the Adjusted Rand Index, Normalized Mutual Information, and silhouette scores to determine the optimal cluster count.

## 3. Results

### 3.1 Phenotype Distribution

Of 177 PCOS-positive patients, the proxy criteria identified 46 (26.0%) as phenotype A, 16 (9.0%) as phenotype B, 36 (20.3%) as phenotype C, and 25 (14.1%) as phenotype D (See **Figure 2**). The remaining 54 (30.5%) did not meet two out of three of the criteria and thus were marked as Unclassified rather than forced into a phenotype category. That nearly one-third of the positive patients fail to clearly map onto any Rotterdam phenotypes through these proxies is a notable finding, underscoring the complexity of clinical presentation.

**Figure 2:**
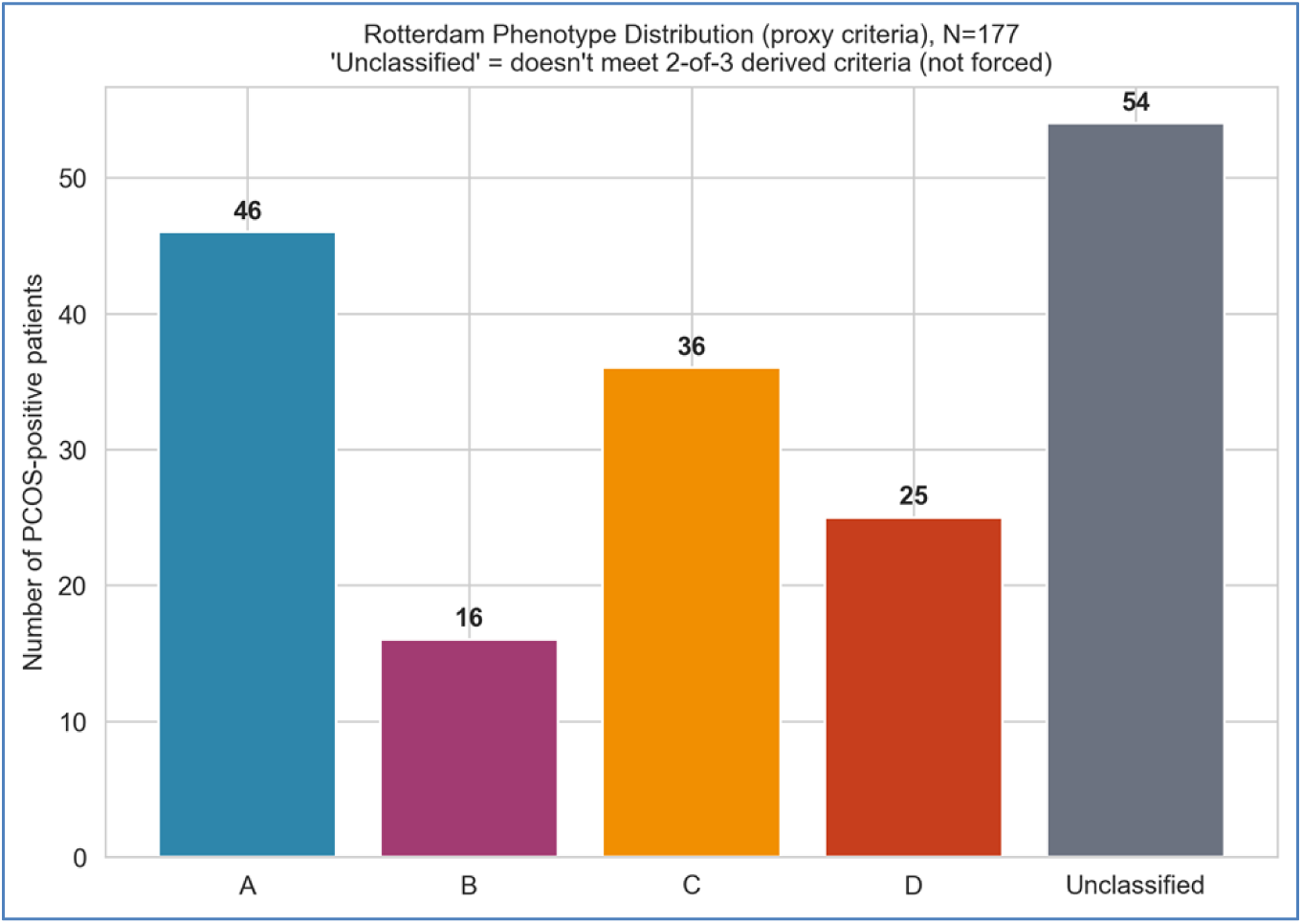
Distribution of Rotterdam phenotypes among PCOS-positive patients (N=177). Phenotype A is the most prevalent (26.0%), while Phenotype B is the rarest (9.0%). The Unclassified group (30.5%) comprises patients who did not meet two out of three proxy criteria.

### 3.2 Binary Benchmark

**Table 3** presents the performance of models trained without tuning, evaluated via ten-fold cross-validation. Random Forest performed best by AUC (0.962), edging out Gradient Boosting (0.961) and XGBoost (0.956).

**Table 3:** Binary classifier benchmark, ten-fold stratified cross-validation, full feature set.

| Model | Accuracy | AUC | F1 | Precision | Recall |
| --- | --- | --- | --- | --- | --- |
| Logistic Regression | 89.3% | 0.946 | 0.833 | 0.850 | 0.820 |
| Random Forest | 91.3% | 0.962 | 0.859 | 0.914 | 0.814 |
| Gradient Boosting | 91.1% | 0.961 | 0.860 | 0.891 | 0.831 |
| SVM (RBF) | 88.5% | 0.943 | 0.814 | 0.870 | 0.768 |
| XGBoost | 90.4% | 0.956 | 0.847 | 0.885 | 0.814 |

Following RandomizedSearchCV tuning, the AUCs of Random Forest and XGBoost improved marginally to 0.963 and 0.961, respectively (**Figure 3**). This small difference indicates the untuned model effectively captures the underlying distribution, confirming that the phenotype gap discussed below is an intrinsic property of the model and not a tuning-induced artifact.

**Figure 3:**
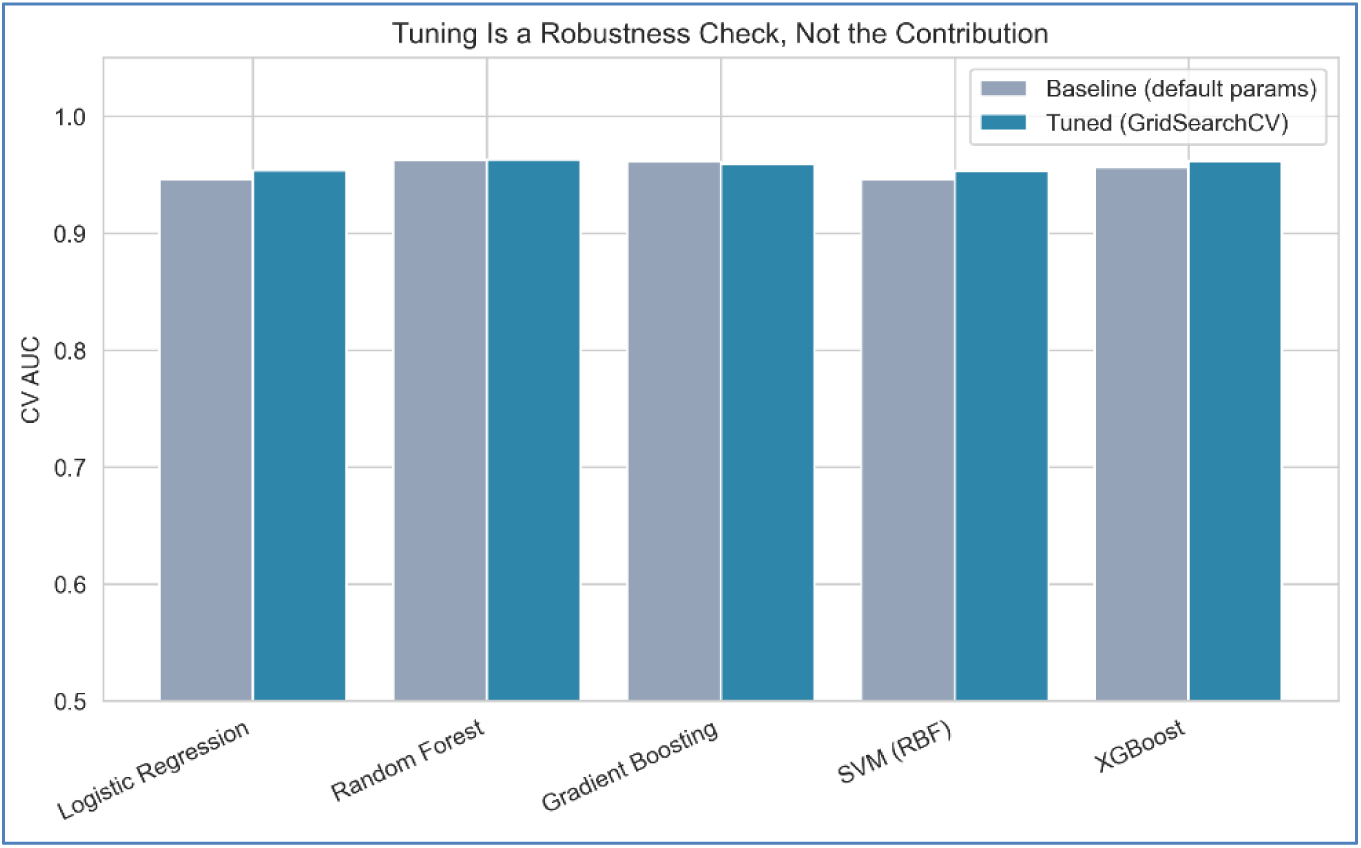
Cross-validated AUC comparison of baseline (untuned) versus tuned classifiers. The marginal improvement after tuning confirms that the baseline models were not substantially under-fit.

Following RandomizedSearchCV tuning, the AUCs of Random Forest and XGBoost improved to 0.963 and 0.961, respectively (**Figure 3**). This marginal improvement confirms that the untuned models were not under-fit—an important confirmation, since the phenotype-level accuracy gap reported below must reflect an intrinsic property of the classification task, not a tuning artifact.

### 3.3 The Core Finding

**Table 4** details the per-phenotype accuracy of the tuned Random Forest on pooled out-of-fold predictions. For the full feature set, accuracy ranged from 75.0% for phenotype B (95% CI 56.2-93.8%) to 97.8% for phenotype A (95% CI 93.5-100%) - a 22.8 percentage-point spread not captured by the overall model’s 90.0% accuracy (See **Figure 4**). Excluding seven phenotype-defining features increased this spread slightly to 23.3 percentage-points; accuracy for each phenotype decreased in accordance with removing the signals from the data, yet the relative spread across phenotypes remained consistent. This is a central empirical finding: the performance gap persists independently of the model’s access to the variables defining the phenotypes.

**Figure 4:**
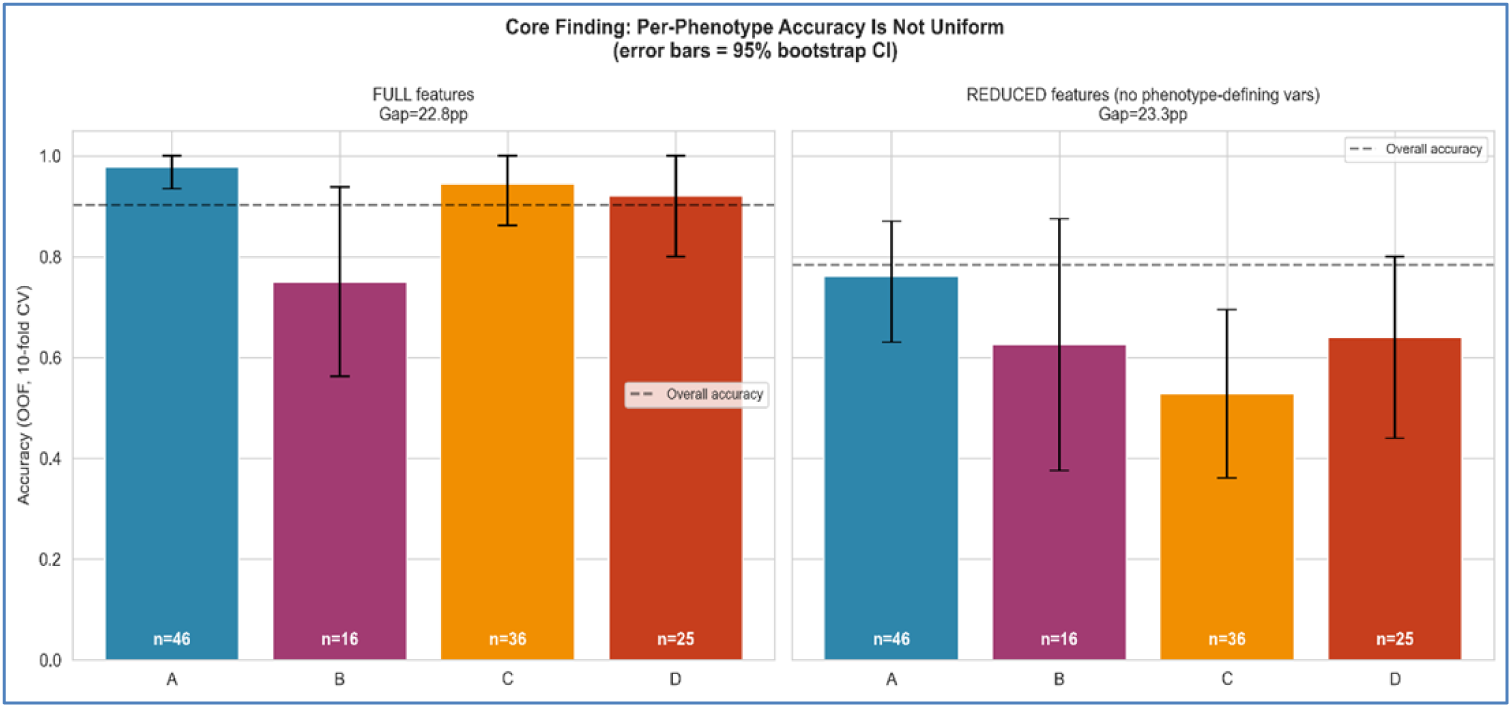
Per-phenotype accuracy with 95% bootstrap confidence intervals for the tuned Random Forest on full and reduced (circularity-controlled) feature sets. The accuracy gap persists even after removing the seven phenotype-defining features.

**Table 4:**
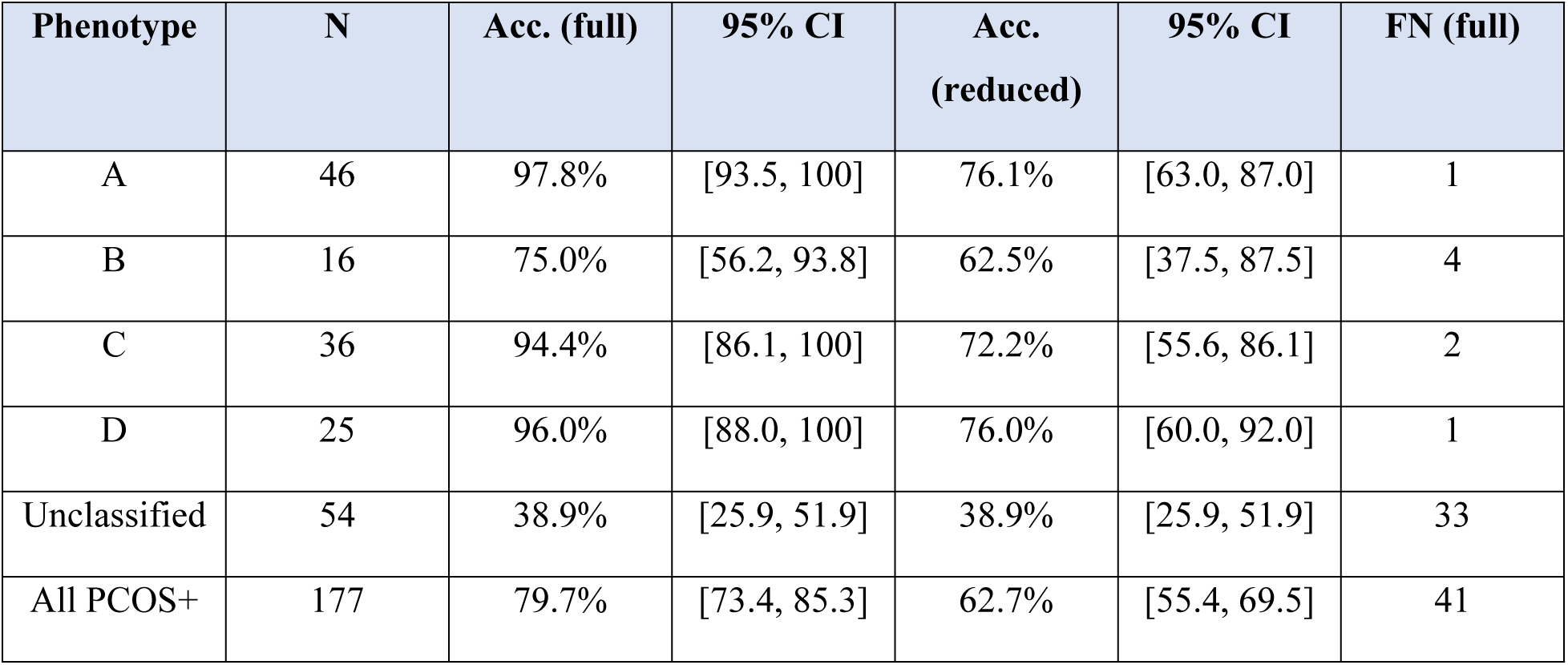
Per-phenotype accuracy of the tuned Random Forest, pooled ten-fold out-of-fold predictions, full versus reduced (circularity-controlled) feature sets.

| Phenotype | N | Acc. (full) | 95% CI | Acc. (reduced) | 95% CI | FN (full) |
| --- | --- | --- | --- | --- | --- | --- |
| A | 46 | 97.8% | [93.5, 100] | 76.1% | [63.0, 87.0] | 1 |
| B | 16 | 75.0% | [56.2, 93.8] | 62.5% | [37.5, 87.5] | 4 |
| C | 36 | 94.4% | [86.1, 100] | 72.2% | [55.6, 86.1] | 2 |
| D | 25 | 96.0% | [88.0, 100] | 76.0% | [60.0, 92.0] | 1 |
| Unclassified | 54 | 38.9% | [25.9, 51.9] | 38.9% | [25.9, 51.9] | 33 |
| All PCOS+ | 177 | 79.7% | [73.4, 85.3] | 62.7% | [55.4, 69.5] | 41 |

The Unclassified group exhibited the lowest accuracy: 33 of its 54 patients were misclassified using the full feature set. The model performs less accurately precisely where the underlying clinical label itself is most ambiguous, reflecting genuine diagnostic complexity.

The Unclassified group shows the lowest accuracy of any category: 33 of its 54 patients are misclassified using the full feature set. The model performs worst where the underlying clinical label is itself most ambiguous—a pattern consistent with genuine diagnostic uncertainty rather than a modeling artifact.

### 3.4 Unsupervised Clustering

Silhouette analysis for k-means clustering identified three clusters as optimal for the PCOS-positive patients. The agreement between the clustering results and the phenotype labels is minimal - the Adjusted Rand Index is 0.045, and the Normalized Mutual Information is 0.052 (where 0 indicates random agreement and 1 indicates perfect agreement) (**Figure 5**). The structure utilized by supervised models diverges from the primary structure discovered by unsupervised approaches on the original feature space. This indicates the supervised signal differs from the dominant unsupervised structure, while confirming the robustness of the accuracy differences identified above.

**Figure 5:**
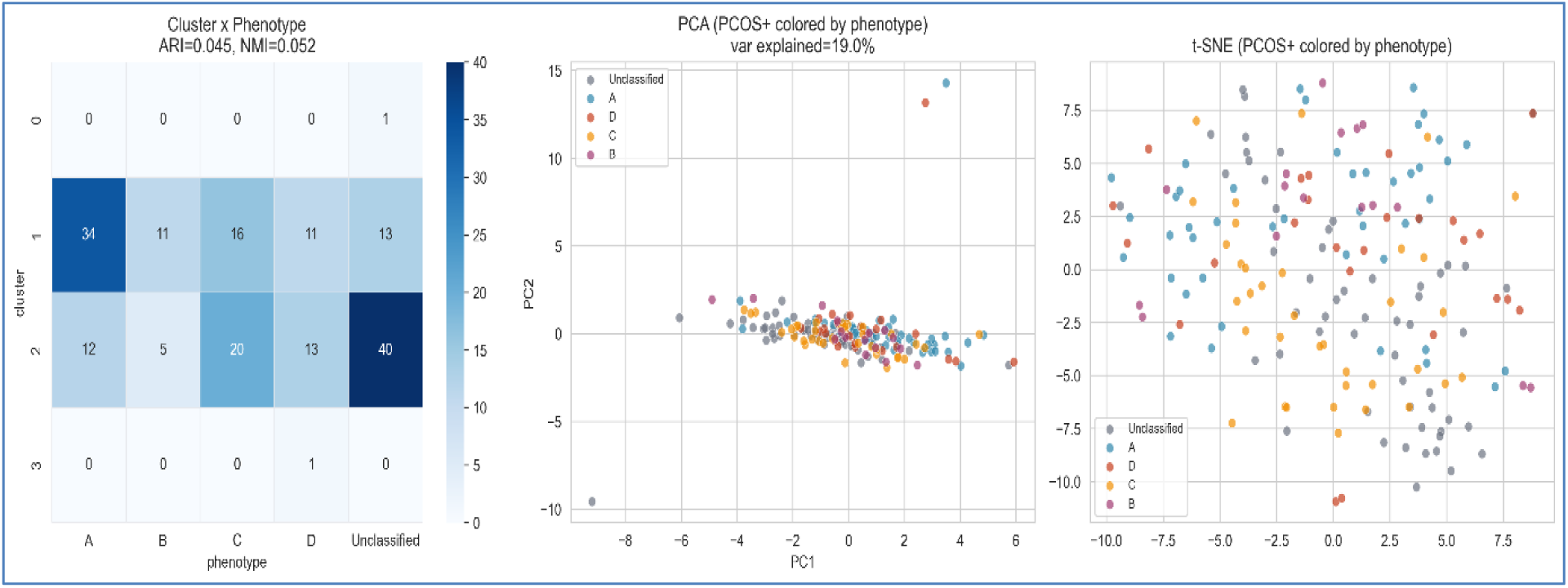
Left: Cluster-by-phenotype cross-tabulation heatmap. Middle: PCA projection of PCOS-positive patients colored by phenotype. Right: t-SNE projection colored by phenotype. The weak alignment (ARI = 0.045) between clusters and phenotypes indicates that supervised and unsupervised structures in the feature space are largely independent.

### 3.5 Multi-Class Trade-Off

Training a class-weighted Random Forest to predict phenotype directly achieved a macro-F1 of 0.661 and an overall accuracy of 83.9%. Accuracy on phenotypes A and D was strong, approaching 100% for each. Given the class imbalance between Control and phenotype B, balanced accuracy (63.2%) provides a clearer summary. **Figure 6** and **Table 5** compare phenotype recall against the original binary classifier: recall for phenotypes A and D improved substantially (100% and 96.0% respectively), but phenotype B’s recall decreased from 75.0% to 43.8%, with nine of its sixteen patients classified directly as Control (**Figure 7**). The Unclassified group’s recall similarly decreased to 31.5%, with 37 of its 54 patients classified as Control.

**Figure 6:**
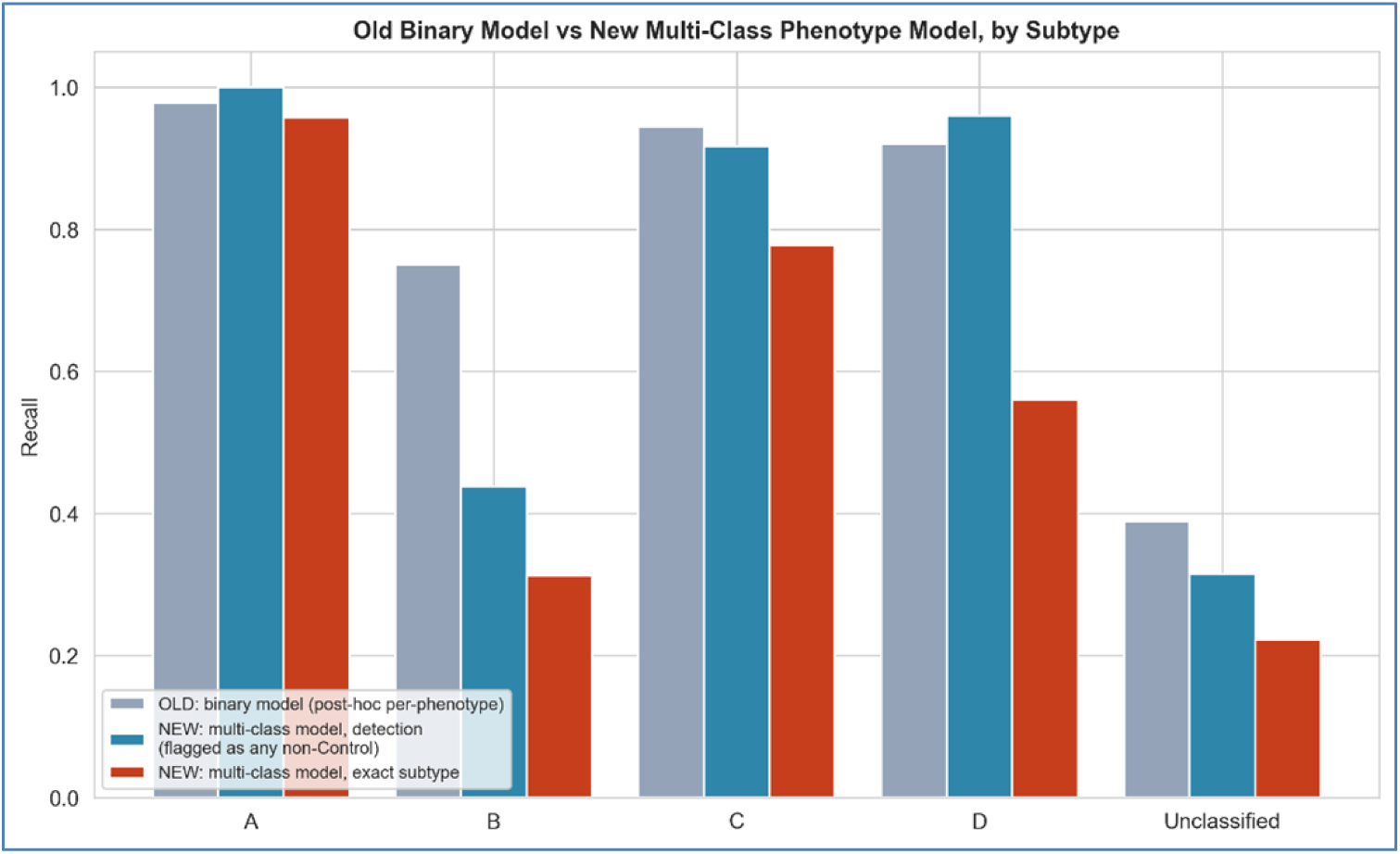
Comparison of detection recall and exact-subtype recall of the multi-class-only model against the original binary classifier, stratified by phenotype.

**Figure 7:**
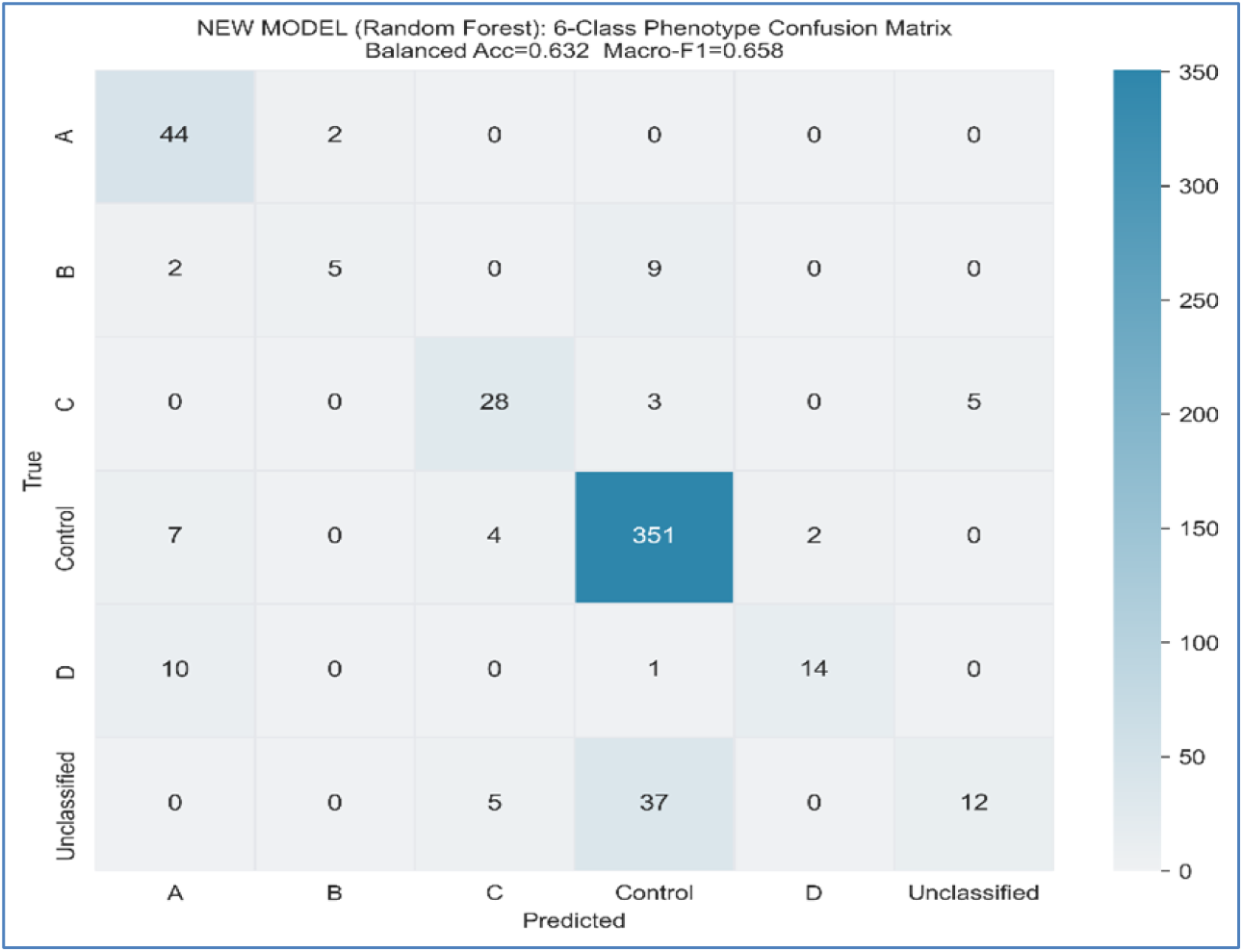
Confusion matrix for the six-class multi-class-only model, based on pooled out-of-fold predictions. Note the concentration of phenotype B and Unclassified misclassifications into the Control class.

**Table 5:** Detection recall comparison: original binary model versus multi-class-only model, by phenotype.

| Phenotype | N | Old (binary)<br>recall | Multi-class-only:<br>detection | Multi-class-only:<br>exact subtype |
| --- | --- | --- | --- | --- |
| A | 46 | 97.8% | 100.0% | 95.7% |
| B | 16 | 75.0% | 43.8% | 31.3% |
| C | 36 | 94.4% | 100.0% | 83.3% |
| D | 25 | 96.0% | 96.0% | 80.0% |
| Unclassified | 54 | 38.9% | 31.5% | 31.5% |

### 3.6 Hybrid Recovery

The genetic algorithm converged on an optimal architecture with 64 nodes in the hidden layer, 0.03 learning rate, equal weighting on the two loss terms, L2 penalty of 0.01, top 15 features selected by Random Forest, and 350 training epochs. Using the same 10-fold out-of-fold evaluation scheme, the hybrid model achieved a binary accuracy of 90.2% and an AUC of 0.962, matching the dedicated binary classifier’s pooled out-of-fold accuracy of 90.2%. Crucially, the hybrid model increased phenotype balanced accuracy to 73.1% and macro F1 score to 0.660, demonstrating significant improvements over the multi-class only model (63.2% and 0.658).

**Table 6** and **Figure 8** detail this comparison, which forms a major contribution to the study. The detection recall of phenotype B increased to 87.5%, exceeding both the original binary model (75.0%) and the multi-class only model (43.8%).

**Figure 8:**
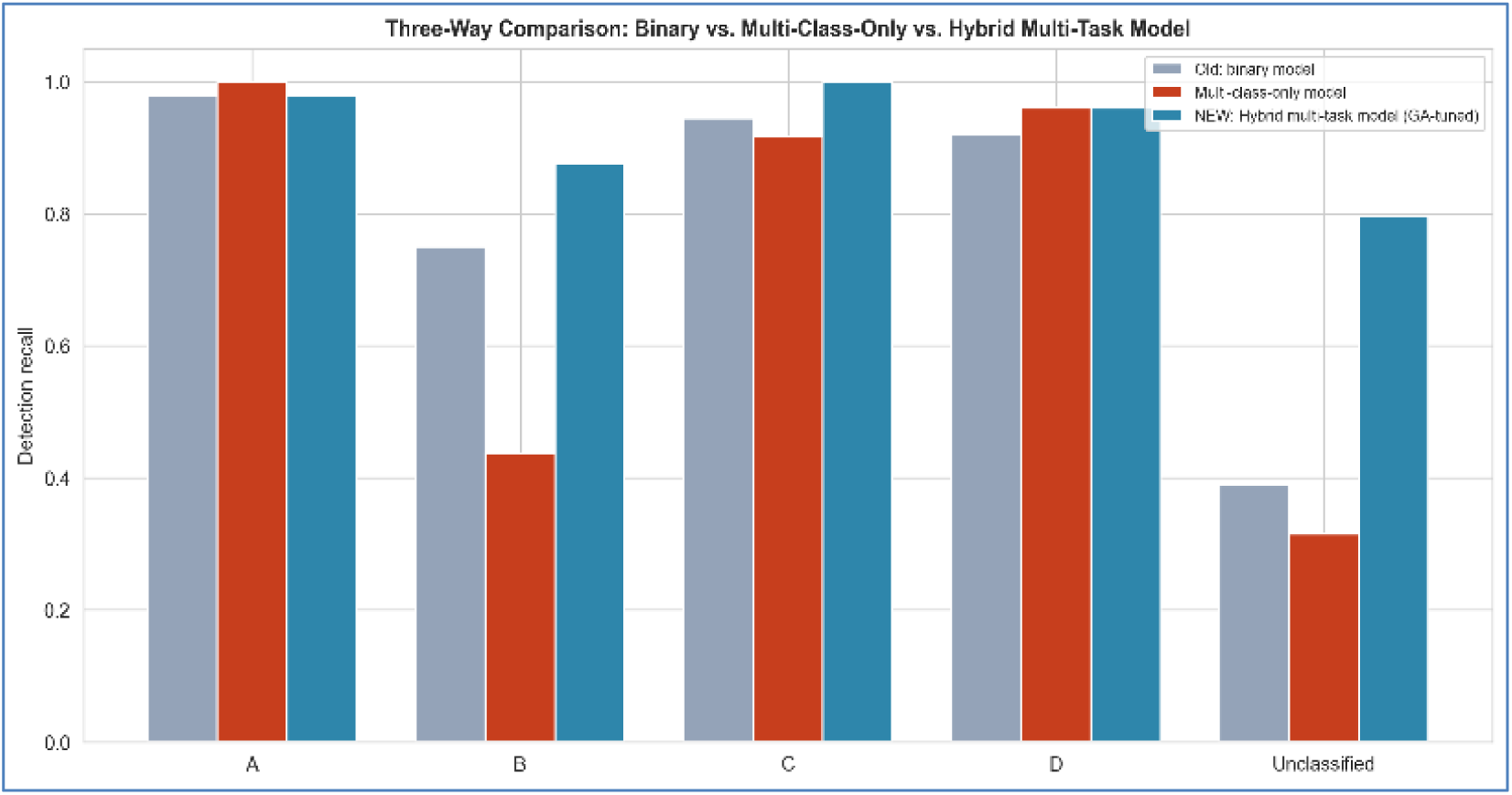
Detection recall by phenotype across the three models: the original binary classifier, the multi-class-only model, and the proposed hybrid multi-task model.

**Table 6:**
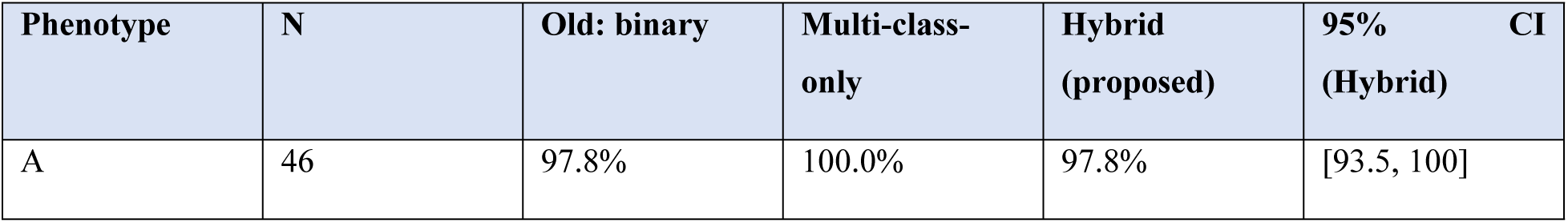
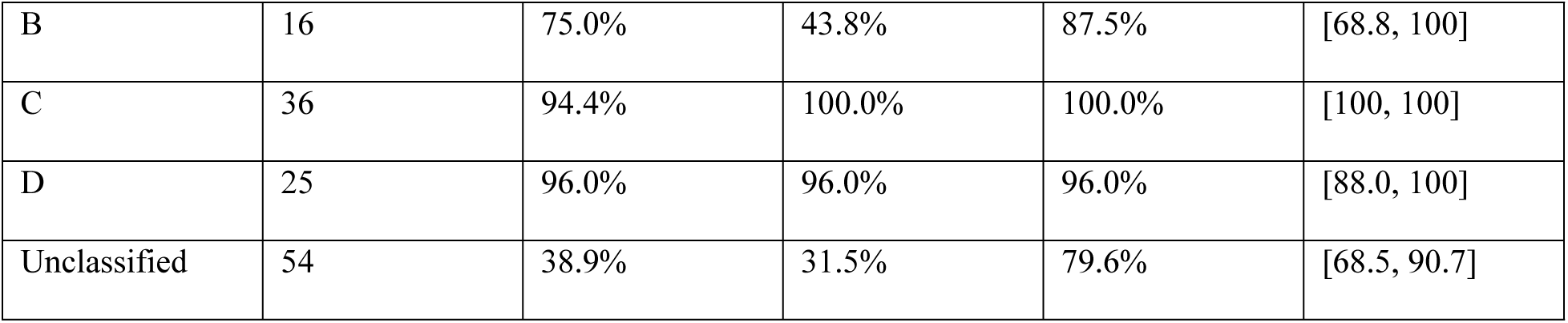
Detection recall by phenotype across all three models, with 95% bootstrap confidence intervals for the hybrid model.

Recall in detection improved to 79.6% for the hybrid model, doubling the value achieved by the multi-class-only model and leaving 11 out of 54 patients classified as Control and 9 as phenotype C. Detection accuracy of phenotype C approached perfection at 100%. Detection accuracy of phenotype D remained stable at 96.0%. The hybrid model achieves this by trading some exact-subtype precision on the majority phenotype A for substantial gains in the rare categories: while detection recall is equal for both models at 97.8%, 33 out of 46 phenotype A patients (71.7%) were classified precisely, compared to the multi-class-only model’s 95.7% accuracy.

These results establish a clear advantage: jointly training the binary and phenotype tasks on selected features, combined with genetic architecture tuning, successfully restores the rare-class performance lost by the multi-class-only model. This represents a highly favorable trade-off, as phenotype B and the Unclassified group are precisely the categories critical for reliable clinical detection.

Paired exact tests (hybrid versus multi-class-only detection, using the same patients; see Section 3.8): phenotype A *p* = 1.00; B *p* = 0.016; C *p* = 0.25; D *p* = 1.00; Unclassified *p* < 0.0001.

**Figure 9** presents the recovery pattern in line-plot format, while **Figure 10** provides the complete six-class confusion matrix from the hybrid model. The line plot makes the recovery pattern visible at a glance: the hybrid model’s detection recall closely tracks the original binary classifier for phenotypes A, C, and D, while rising substantially above both alternatives for phenotypes B and Unclassified—the two subgroups a clinician would least want a screening tool to miss.

**Figure 9:**
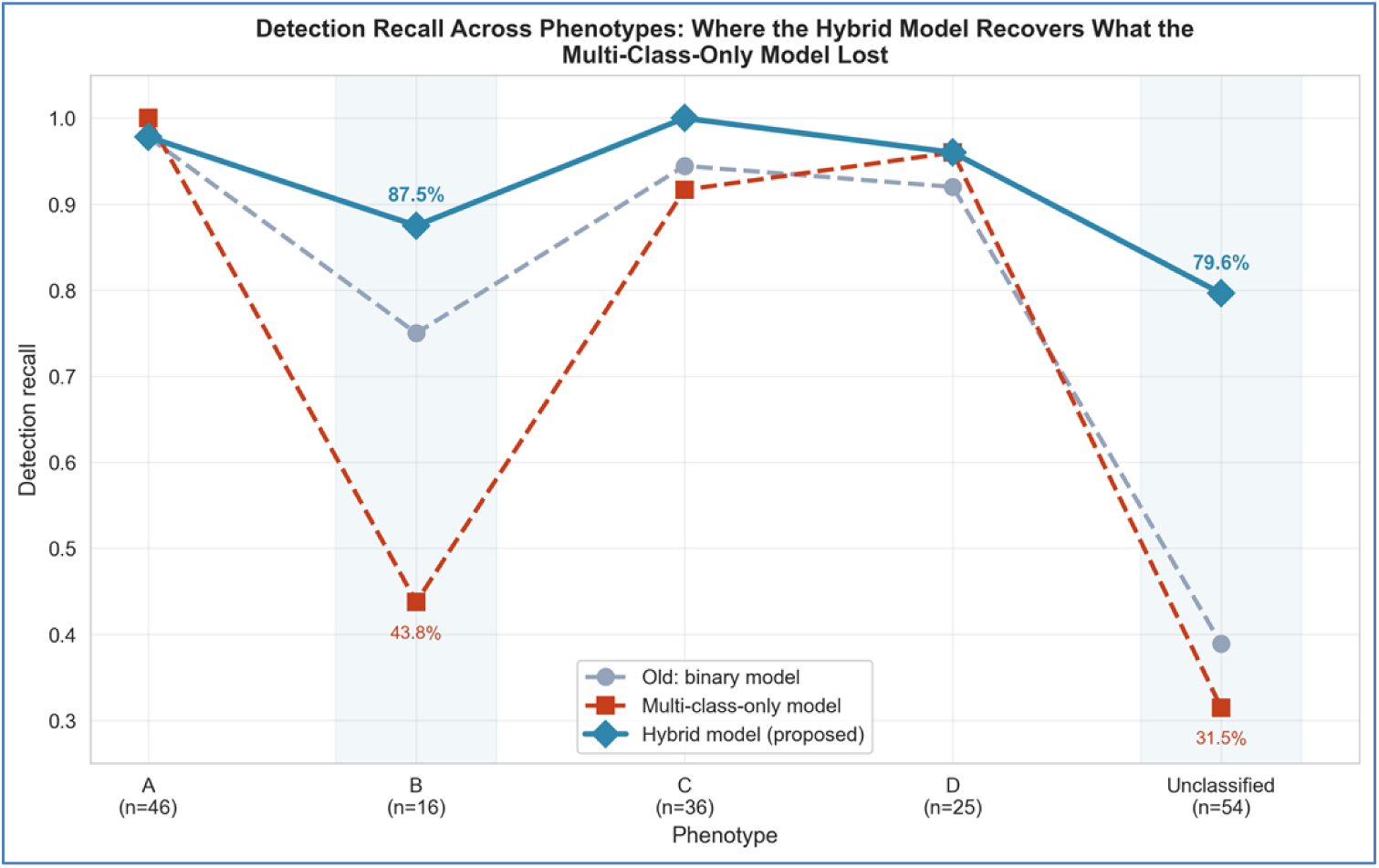
Line plot of detection recall by phenotype, illustrating the recovery pattern achieved by the hybrid model relative to the multi-class-only baseline.

**Figure 10:**
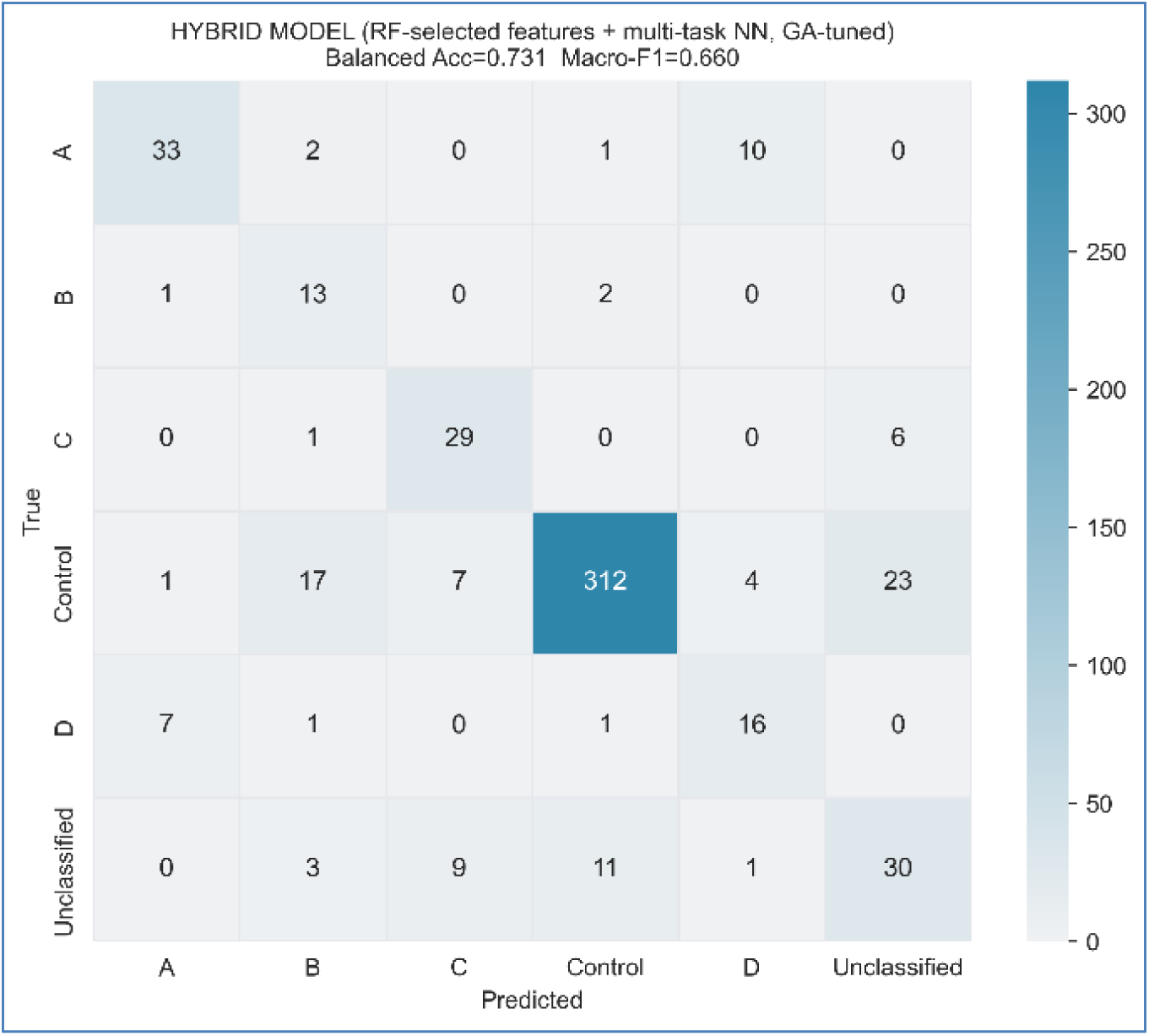
Six-class confusion matrix for the hybrid multi-task model, based on pooled out-of-fold predictions.

In the case of Unclassified cases, detection recall rose up to 79.6%, which is more than double than the performance achieved with multi-class-only classifier. In the group of 54 Unclassified cases, 11 were found to be Control and 9 phenotype C. The detection rate for phenotype C was 100%, and the one for phenotype D remained 96.0%. However, the one flaw of the hybrid model is the exact-subtype precision for phenotype A. Even though the detection recall remains at 97.8%, only 33 of 46 patients (71.7%) had the right label, compared with 95.7% under multi-class-only model with 10 phenotype-A patients mislabeled as phenotype D. On a clinical standpoint, the tradeoff is favorable for the hybrid model, as the improvement in detection of the rarest and hardest-to-diagnose phenotypes comes at the expense of mislabeling of the phenotype A patients as phenotype D in the subphenotype assignment of the most common phenotypic subgroup of PCOS.

### 3.7 Independent Reproduction

The complete approach has been replicated independently on the second data set with 541 patients. This independent replication delivered identical results: 90.2% pooled out-of-fold binary accuracy, 22.8 percentage points gain on the full feature set (23.3 on the reduced one) and adjusted Rand index (ARI) equal to 0.045 with the same exact solution found by the genetic algorithm. This indicates that the whole approach is fully deterministic and replicable.Due to the absence of similarly comprehensive public datasets containing both necessary hormonal measurements and follicle counts, a covariate shift stress test was conducted to assess stability. All three models were tested on a perturbed dataset simulating measurement drift (10-35% of each feature’s SD and an 8% random flip rate of binary clinical signs). None of the models experienced performance degradation, confirming their stability under shifted data distributions (**Figure 11**).

**Figure 11:**
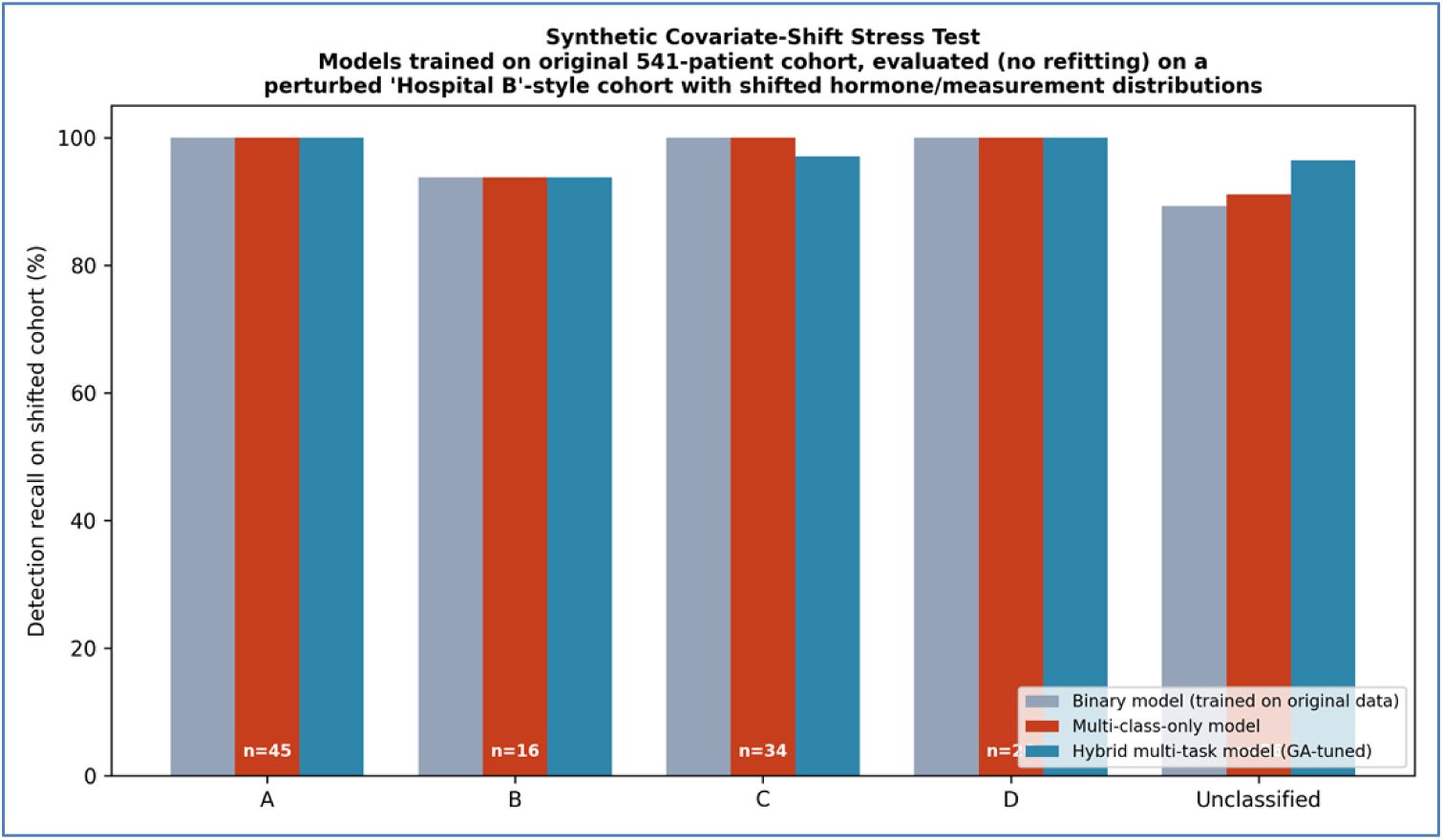
Synthetic covariate-shift stress test. Detection recall by phenotype for all three models, trained on the original 541-patient cohort and evaluated on a perturbed cohort simulating inter-hospital measurement drift (10–35% feature-level noise, 8% binary-sign flip rate).

### 3.8 Statistical Comparison and Ablation

A McNemar’s test comparing the Random Forest and hybrid models on the binary task showed no significant difference in classification (p = 1.00, with 19 patients correct only by the former and 19 only by the latter).

**Table 7** presents a paired exact test for detection recall between the hybrid and multi-class-only models. The improvements are highly statistically significant: for phenotype B, seven patients were detected by the hybrid model but not by the multi-class model, with zero in reverse (p = 0.016, bootstrap CI [+18.8, +68.8]pp). For the Unclassified group, 26 patients were detected only by the hybrid model, and 0 only by the multi-class model (p < 0.0001). There was no significant difference between A and D (p = 1.00), or C (p = 0.25).

**Table 7:**
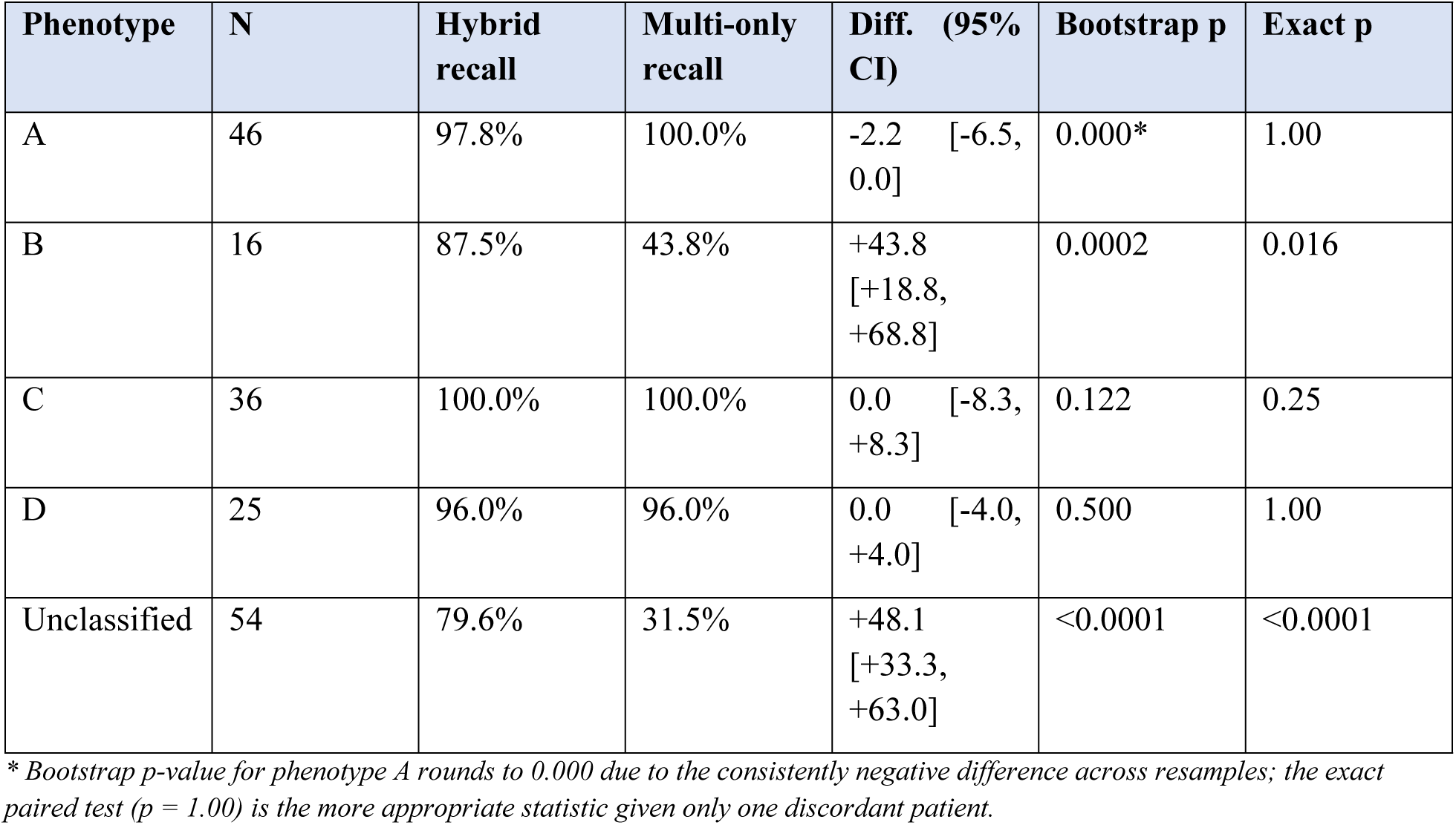
Paired significance tests: detection recall of the hybrid versus multi-class-only model on the same out-of-fold predictions.

| Phenotype | N | Hybrid recall | Multi-only recall | Diff. (95% CI) | Bootstrap p | Exact p |
| --- | --- | --- | --- | --- | --- | --- |
| A | 46 | 97.8% | 100.0% | -2.2 [-6.5, 0.0] | 0.000* | 1.00 |
| B | 16 | 87.5% | 43.8% | +43.8 [+18.8, +68.8] | 0.0002 | 0.016 |
| C | 36 | 100.0% | 100.0% | 0.0 [-8.3, +8.3] | 0.122 | 0.25 |
| D | 25 | 96.0% | 96.0% | 0.0 [-4.0, +4.0] | 0.500 | 1.00 |
| Unclassified | 54 | 79.6% | 31.5% | +48.1 [+33.3, +63.0] | <0.0001 | <0.0001 |

\* Bootstrap p-value for phenotype A rounds to 0.000 due to the consistently negative difference across resamples; the exact paired test ( $p = 1.00$ ) is the more appropriate statistic given only one discordant patient.

The ablation study performed in **Table 8** provides interesting insights into the architectural factors responsible for the improved results. The variant with only phenotype achieved 71.7% balanced accuracy, 0.659 macro-F1, 87.5% phenotype B recall, and 77.8% Un-classified recall with results being almost identical to those of the full hybrid network. Moreover, the variant without a shared layer was able to produce 89.8% binary accuracy, 0.956 AUC, 93.8% phenotype B recall, and 81.5% Unclassified recall.

**Table 8:**
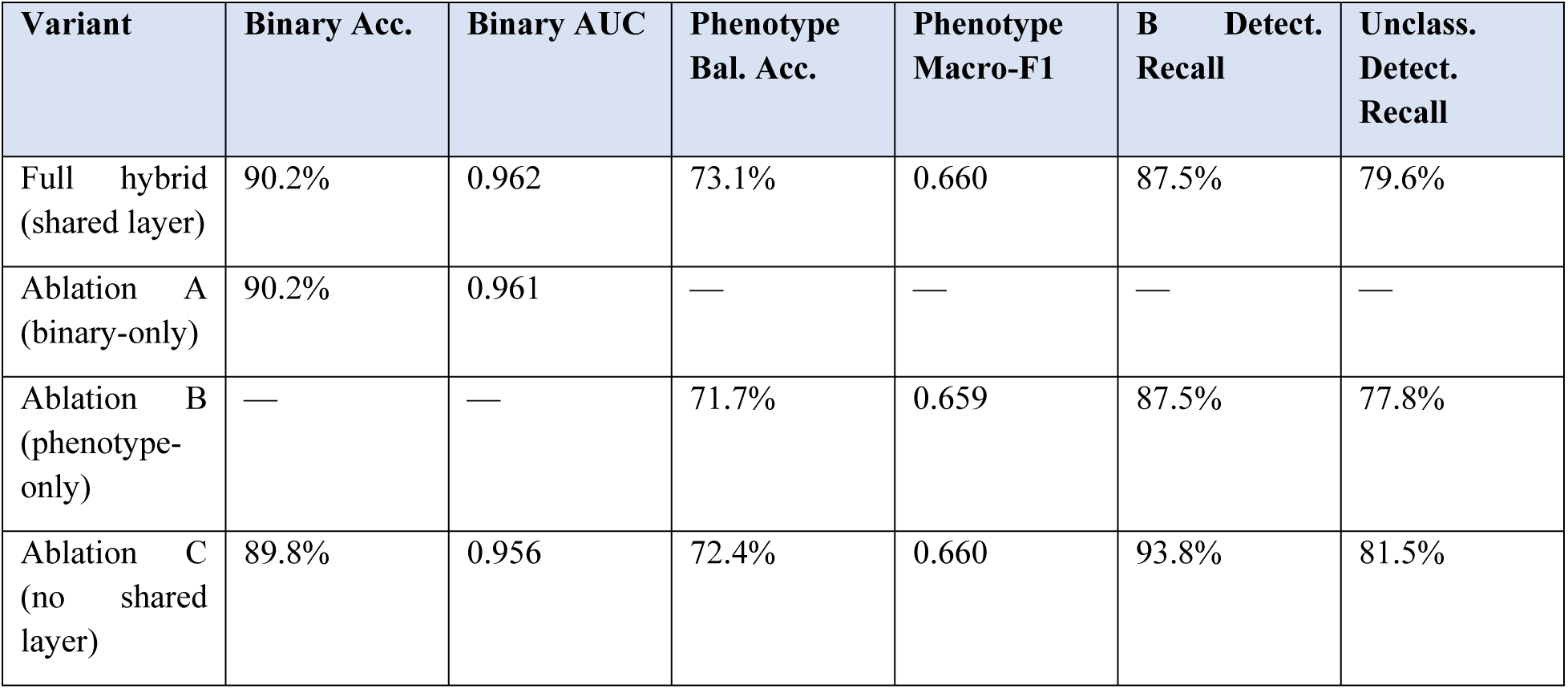
Ablation results. All variants use the GA-selected hyperparameters, the top 15 Random-Forest-ranked features, and the same ten-fold out-of-fold splits as the full hybrid model.

From the ablation study, one can gain better understanding about the architectural factors responsible for the improvement, namely feature selection and class weighting. In turn, this allows for more practical use of the hybrid model as the shared layer architecture is not a necessity for achieving the improvements.

Binary model (binary-only) showed 90.2% accuracy and 0.961 AUC, performing identically to the full hybrid model and suggesting that removing the phenotype head does not harm the binary model. The phenotype-only model (ablation B) resulted in 71.7% balanced accuracy, 0.659 macro-F1 score, 87.5% phenotype-B recall and 77.8% Unclassified recall, being already quite close to the full hybrid. The ablation C, which used no shared layer and had two independently trained heads optimized using sum of their losses, showed 89.8% binary accuracy, 0.956 AUC, 93.8% phenotype-B recall and 81.5% Unclassified recall and thus was either comparable or superior to the full hybrid in terms of phenotype recovery. The 5-seed stability test confirms this tendency: the no-shared-layer version shows 92.5% (std 4.7) phenotype-B recall against 90.0% (std 5.0) of the shared one, with Unclassified recall not being statistically different (81.1% vs 81.5%), while binary accuracy is consistently somewhat worse (89.8% vs 90.1%).

Ablation shows that it is the architectural components involved in the recovery process which lead to the good recovery performance. The 15-feature reduction through Random Forest and the class weighting through inverse frequency in the phenotype loss function are the two main reasons why recovery performances on phenotypes B and Unclassified are improved. Even though the shared hidden layer is not required for the recovery of rare phenotypes, there is a benefit from having it as a single trained model can now do both the binary and the phenotype classification tasks at once, thus avoiding the necessity of training and using two separate models. The value of this study is the whole joint training process including the feature selection, class-balanced loss, and multi-task network rather than the representation sharing as a stand-alone approach.

## 4. Discussion

There are three main facts that together make the argument for phenotype-aware evaluation of PCOS machine learning models. The 22.8 percentage point gap in accuracy among the phenotypes A through D clearly shows that the overall number of 90% accuracy can mask the fact of how well or bad the model performs on individual patients. This reveals a significant blind spot shared by prior approaches, including the otherwise strong binary performance validation presented by Nasim et al. (2022) [6], Rahman et al. (2024) [9], and Agirsoy and Oehlschlaeger (2025) [8].

The maintenance of the gap after the elimination of variables used in the definition of the phenotype serves as an important guard against circularity, as it rules out the possibility that the noted difference has been caused by variables that are mere correlates of the clinical criteria excluded. While this may not necessarily mean that there is a biological difference between the two phenotypes, it does show that the classifiers are using secondary variables that do not equally reflect the sub-groups. Prior work, such as Zad et al. (2024) [5], labeled PCOS patients based on Rotterdam criteria components without verifying the consistency of their modeling after splitting patients by their criterion fulfillment. This work provides that necessary verification.

The inability of the normal multi-class classifier to recognize phenotype B shows that simply changing the classification target is not enough. The increase in the number of classes requires more training data in each class, which rare cases like phenotype B don’t have. While Silva et al. (2022) [3] used feature selection and clustering to explore patient subgroups, they did not evaluate the variation of classifier accuracy between the resultant classes. Thus, this work proves that phenotype-aware modeling does not necessarily yield better results compared to binary classification when rare phenotypes are not well represented, addressing a significant limitation of existing research.

To overcome the aforementioned limitation, a joint training paradigm is used in this study which allows recovering the performance on rare subgroups. The hybrid classifier combines tree-based feature selection and a neural component and uses hyperparameters tuned using genetic algorithm optimization. Even though similar methods have already been employed to build models for PCOS datasets-Such as the genetic algorithm tuning by Ghosh and Srinivasan (2025) [12] or the ensemble approaches by Faris et al. (2025) [13] and Pallavi and Soumya (2026) [11]— those approaches usually try to predict a binary or image-level outcome. By predicting the binary outcome and the phenotype together, our model lifts phenotype B’s recall from 43.8% to 87.5% and the Unclassified group’s recall from 31.5% to 79.6%, while preserving binary classification accuracy. Ablation study shows that such improvement was achieved through targeted feature selection and balanced training, making deployment feasible.

Furthermore, weak clustering-phenotype alignment (ARI = 0.045, NMI = 0.052) indicates that the differences between phenotypes do not form the main structure of the data. Such result is a natural consequence given the overlapping nature of the Rotterdam phenotypes, which are based on three criteria only. Nevertheless, weak alignment of clusters and phenotypes does not exclude the existence of the accuracy gap, but rather highlights the possibility of exploiting structures missed by the unsupervised approach.

### 4.1 Clinical Relevance and the Case for Phenotype-Aware Screening

The subgroup gap documented here is not only a modeling curiosity; it tracks a real, previously established clinical difference. In a retrospective cohort of 258 PCOS patients and 110 controls, Shroff et al. (2007) [19] found that three of the four Rotterdam phenotypes carried a significantly elevated risk of metabolic syndrome relative to controls - age-adjusted odds ratios of 6.3 to 8.2 - while the fourth did not differ from controls at all. In a larger, 2,288-patient multicenter cohort, Daan et al. (2014) [20] likewise found that cardiometabolic risk factor prevalence differed meaningfully across phenotypes, concentrated in the hyperandrogenic subtypes. Phenotype B - the specific subgroup this paper’s binary classifier detects worst (75.0% recall) and the hybrid model recovers most (87.5% recall) - is exactly this kind of hyperandrogenic, oligo-ovulatory phenotype. A classifier that silently under-detects phenotype B is not failing at a label of purely academic interest; on the evidence above, it is failing at identifying patients in a higher metabolic-risk category who would benefit most from early screening and counseling.

This is also precisely why the absence of prospective clinical evaluation, flagged as the central concern in this paper’s review, is treated as a first-order limitation rather than a closing remark. Everything reported here is a retrospective analysis of a single public benchmark with proxy phenotype labels; no claim is made, or should be inferred, that the hybrid model is ready for clinical deployment, and none of its numbers constitute evidence of clinical benefit in a real diagnostic pathway. What this paper establishes is narrower and, we think, still useful: that the phenotype-level gap is real, that it lines up with a clinically meaningful risk distinction documented independently in the endocrinology literature, and that a specific, interpretable modeling change closes most of it on the data available. Section 7 lays out concretely what would be required to move from that finding to a claim about clinical utility - a biochemically confirmed cohort, a paired statistical comparison against current screening practice, and eventually a prospective decision-impact study - and none of those steps has been taken yet.

The strongest available evidence that this general premise is correct arrived after the studies above: Gao et al. (2025) [21], published in Nature Medicine, identified four data-driven PCOS subtypes by unsupervised clustering of 9 clinical variables in 11,908 women, validated across five international cohorts (China, USA, Europe, Singapore, Brazil), and followed a subset prospectively for a median of 6.5 years. The subtypes are defined differently from the four Rotterdam phenotypes analyzed in this paper - by clustering continuous hormonal and metabolic variables rather than by combinatorial Rotterdam criteria - but the clinical conclusion is the same in kind: subtype membership predicted materially different outcomes, including second-trimester pregnancy loss risk (odds ratio 7.32 for the hyperandrogenic subtype), live birth rates after IVF ranging from 48.9% to 56.3% across subtypes, and markedly different rates of type 2 diabetes, hypertension, and ovarian hyperstimulation syndrome. This is, to our knowledge, the largest and most rigorously validated real-world demonstration to date that PCOS subtype is not a statistical artifact but a determinant of clinically material outcomes at a scale (multi-country, prospectively followed, over 11,000 patients) this paper’s retrospective, single-benchmark analysis cannot approach. It is offered here not as validation of this paper’s specific hybrid model - it is not - but as independent, large-scale clinical grounding for the premise that motivates it: that collapsing PCOS heterogeneity into a single binary label discards information a patient’s long-term health outcome actually depends on.

## 5. Limitations

The most consequential limitation of this work is stated first rather than buried: every result in this paper comes from a single, secondary analysis of one public, retrospective benchmark dataset, with no prospective data collection, no biochemically confirmed diagnosis, and no clinical evaluation of any kind. Nothing here has been tested on an independent cohort, reviewed by a treating clinician, or evaluated for whether it changes a real diagnostic or management decision. The statistical comparisons in Section 3.8, the ablation study, and the clinical-relevance argument made in Section 4.1 strengthen the internal validity of the phenotype-gap finding and the hybrid model’s recovery of it, but internal validity on a public benchmark is not clinical evidence, and this paper does not claim otherwise. Sections 4.1 and 7 lay out specifically what a credible clinical evaluation would require and why none of it has been done yet.

The primary limitation of this work relates to the available data. The dataset lacks biochemical androgen measurements, which are standard for the hyperandrogenism component of the Rotterdam criteria. Consequently, the phenotype labels depend on a proxy derived from hirsutism, acne, and relative hormonal levels, a constraint similarly encountered by Zad et al. (2024) [5]. The sample consists of 541 subjects who come from a single state in India, which limits the applicability of the results. Phenotype B has just sixteen subjects in its sample, and while the use of out-of-fold validation helps eliminate bias, the confidence intervals for performance on the subsample are still quite wide. Internal thresholds and the Unclassified status for nearly one-third of positive cases are to be considered an artifact of the dataset rather than epidemiological reality.

Several limitations have been placed on methodology intentionally to keep the focus of the analysis. The hyperparameter tuning has used fixed-budget searching, where twenty iterations of randomized search are performed for baseline models, while the genetic algorithm uses a small population for five generations; wider budgets can cause marginal changes to performance. In addition, the ablation study proves that gains of the hybrid architecture for rare phenotypes happen because of Random-Forest feature selection and phenotype loss function with class weights, not because of the proposed shared hidden representation architecture.

## 6. Conclusion

This work effectively demonstrates that standard machine learning models for PCOS diagnosis hide significant differences in performance across clinical subgroups, and we propose a robust solution to address this problem. Three main contributions of this study are outlined. At first, it exposes a large accuracy gap of 22.8 percentage points between the best and worst performing Rotterdam phenotypes on conventional binary classification, a difference entirely hidden by aggregate reporting metrics. Secondly, it illustrates the multi-class trade-off: training a model to predict phenotypes shifts the failure threshold, with severe consequences for detecting rare manifestations such as phenotype B. Third, it proposes a hybrid training approach that improves performance on rare and unclassified phenotypes, while maintaining overall binary accuracy, as validated statistically. The ablation clearly emphasizes feature selection and class-balanced training as critical elements, offering a streamlined approach for clinical implementation.

While prior models—including optimized approaches like those of Ghosh and Srinivasan (2025) [12] and Faris et al. (2025) [13]—have achieved high aggregate accuracy, this research emphasizes that architectural complexity does not guarantee consistent performance across subgroups. Moving forward, phenotype-aware evaluation should become the standard requirement for evaluating PCOS machine learning models to ensure equitable and reliable clinical applications.

## 7. Future Scope

Two items require no new data collection and are the immediate next step: an extensive seed-and imbalance-sensitivity analysis to quantify how much of the hybrid model’s rare-phenotype recovery depends on the specific random seed used throughout this paper, and a mechanistic ablation that isolates the shared hidden representation’s contribution from the Random-Forest feature selection and class-weighted loss already shown to drive most of the recovery in Section 3.8.

The central, and by far the most important, next step is clinical rather than computational: external validation on a cohort with biochemically confirmed Rotterdam phenotypes - direct androgen assays rather than the proxy used here - ideally collected across multiple sites to also address the single-population limitation. That validation should be designed from the outset around a paired statistical comparison against current binary screening practice, using the same tools applied in Section 3.8 (McNemar’s test, bootstrap confidence intervals on per-phenotype recall), together with a decision-analytic evaluation - reporting where any change in phenotype-B or Unclassified detection translates to a net clinical benefit rather than only a metric improvement.

Only once that step is complete would a clinician-in-the-loop pilot - comparing phenotype-aware flags against standard binary screening on real referral decisions, with clinician review of disagreements - be appropriate, followed eventually by a prospective study of whether phenotype-informed screening changes diagnostic timelines or downstream metabolic outcomes relative to current practice. Separately, the collaborative training method proposed here can be extended to the other genetically optimized or hybrid architectures reviewed in Section 1.1 [15–17], to test whether the joint binary-and-phenotype objective generalizes beyond the specific tree-plus-neural-network design used in this paper.

## Ethics approval and consent to participate

Not applicable.

## Consent for publication

Not applicable.

## Conflicts of Interest

The authors declare no conflicts of interest.

## Data Availability Statement

This study performed a secondary analysis of the publicly available, de-identified Kerala PCOS dataset (https://www.kaggle.com/datasets/prasoonkottarathil/polycystic-ovary-syndrome-pcos/data) (Kottarathil, 2020) [14], collected across ten hospitals in India; no new patient data or identifiers were involved. The Rotterdam phenotype assignments used here are proxy labels derived from existing features, not clinical diagnoses, and should not guide individual patient care. The dataset, phenotype derivation code, model implementations, and scripts for reproducing all results are provided in the accompanying notebook.

## Authors’ Contributions

P.R., S.D. and K.S. participated in conceptualization, method design, data collection and data curation, experiment, writing the manuscript, while P.R. and S.M. validated the results, supervised, reviewed and edited the manuscript.

## Funding

The authors received no funding from their institutes for this study.

## Acknowledgment

We sincerely thank all research team members for the support provided during the conduct of this study.

## Declaration of AI Assistant

The authors have not used AI for any text generation. An AI assistant has been used for language improvement in the result section.

## Notes

### Competing Interest Statement

The authors have declared no competing interest.

## References

[1] Joo, Yoonjung Yoonie, et al. “A polygenic and phenotypic risk prediction for polycystic ovary syndrome evaluated by phenome-wide association studies.” The Journal of Clinical Endocrinology & Metabolism 105.6 (2020): 1918–1936.

[2] Memon, Muzmmil, et al. “Machine Learning Models for Prediction and Management of Polycystic Ovary Syndrome: A Systematic Review.” The Critical Review of Social Sciences Studies 3.4 (2025): 3089–3105.

[3] Silva, I. S., et al. “Polycystic ovary syndrome: clinical and laboratory variables related to new phenotypes using machine-learning models.” Journal of Endocrinological Investigation 45.3 (2022): 497–505.

[4] Ghaderzadeh, Mustafa, Ali Garavand, and Cirruse Salehnasab. “Artificial intelligence in polycystic ovary syndrome: A systematic review of diagnostic and predictive applications.” BMC Medical Informatics and Decision Making 25.1 (2025): 427.

[5] Zad, Zahra, et al. “Predicting polycystic ovary syndrome with machine learning algorithms from electronic health records.” Frontiers in Endocrinology 15 (2024): 1298628.

[6] Nasim, Shazia, et al. “A novel approach for polycystic ovary syndrome prediction using machine learning in bioinformatics.” IEEE Access 10 (2022): 97610–97624.

[7] Govindharajan, Girija, et al. “Innovative AI-driven models for predicting polycystic ovarian syndrome: An extensive review of machine learning and deep learning frameworks.” Archives of Computational Methods in Engineering 33.2 (2026): 2115–2140.

[8] Agirsoy, Mehtap, and Matthew A. Oehlschlaeger. “A machine learning approach for non-invasive PCOS diagnosis from ultrasound and clinical features.” Scientific Reports 15.1 (2025): 33638.

[9] Rahman, Md Mahbubur, et al. “Empowering early detection: A web-based machine learning approach for PCOS prediction.” Informatics in Medicine Unlocked 47 (2024): 101500.

[10] Joyti, Sadia Akther, et al. “Machine Learning and Computational Intelligence Approaches for Prediction and Management of Polycystic Ovary Syndrome.” 2026 IEEE 2nd International Conference on Quantum Photonics, Artificial Intelligence & Networking (QPAIN). IEEE, 2026.

[11] Pallavi, Mrs, and S. Soumya. “An Intelligent Multi-Modal Bi-Gan And Pinn Framework For Early Prediction Of PCOS And Metabolic Syndrome.” International Journal of Advances in Signal and Image Sciences (2026): 238–259.

[12] Ghosh, Ananya, and Kathiravan Srinivasan. “EffiDenseGenOp: Ensemble Transfer Learning with hyperparameter tuning using genetic algorithm optimization for PCOS detection from ultrasound sonography images.” IEEE Access (2025).

[13] Faris, Najlaa, et al. “Enhanced Polycystic Ovary Syndrome diagnosis model leveraging a K-means based genetic algorithm and ensemble approach.” Intelligence-Based Medicine 11 (2025): 100253.

[14] Kottarathil, Prasoon. “Polycystic Ovary Syndrome (PCOS).” Kaggle Dataset, 2020, https://www.kaggle.com/prasoonkottarathil/polycystic-ovary-syndrome-pcos. (access date: 15 January, 2026).

[15] Tasnim, Mayesha, et al. “PCOS detection from ovarian ultrasound images using feature fusion and hybrid classification approaches.” Health Information Science and Systems 14.1 (2026): 75.

[16] Smithakranthi, A., and D. Haritha. “An automated PCOS prediction model using hybrid text and gate network model using learning approaches.” International Journal of System Assurance Engineering and Management (2026).

[17] Kermanshahchi, Julia, et al. “Development of a machine learning-based model for accurate detection and classification of polycystic ovary syndrome on pelvic ultrasound.” Cureus 16.7 (2024).

[18] Teede, Helena J., et al. “Polyendocrine metabolic ovarian syndrome, the new name for polycystic ovary syndrome: a multistep global consensus process.” The Lancet 407.10545 (2026): 2329–2339.

[19] Shroff, Rupal, et al. “Risk of metabolic complications in the new PCOS phenotypes based on the Rotterdam criteria.” Fertility and Sterility 88.5 (2007): 1389–1395.

[20] Daan, Nadine M., et al. “Cardiovascular and metabolic profiles amongst different polycystic ovary syndrome phenotypes: who is really at risk?” Fertility and Sterility 102.5 (2014): 1444–1451.

[21] Gao, Xueying, et al. “Data-driven subtypes of polycystic ovary syndrome and their association with clinical outcomes.” Nature Medicine 31 (2025): 4214–4224.

